# Online prediction of optimal deep brain stimulation contacts from local field potentials in chronically-implanted patients with Parkinson’s disease

**DOI:** 10.1101/2024.11.26.24317968

**Authors:** Marjolein Muller, Stefano Scafa, Ibrahem Hanafi, Camille Varescon, Chiara Palmisano, Saskia van der Gaag, Rodi Zutt, Niels A van der Gaag, Carel F.E. Hoffmann, Jocelyne Bloch, Mayte Castro Jiménez, Julien F. Bally, Philipp Capetian, Ioannis U. Isaias, Eduardo M. Moraud, M. Fiorella Contarino

**Affiliations:** Department of Neurology, Leiden University Medical Center, Leiden, the Netherlands; Department of Clinical Neurosciences, University Hospital Lausanne (CHUV), Lausanne, Switzerland; Defitech Center for Interventional Neurotherapies (NeuroRestore), University Hospital Lausanne and Ecole Polytechnique Fédérale de Lausanne, Switzerland; Institute of Digital Technologies for Personalized Healthcare (MeDiTech), University of Applied Sciences and Arts of Southern Switzerland (SUPSI), Lugano, Switzerland; Department of Neurology, University Hospital of Wuerzburg and Julius Maximilian University of Wuerzburg, Wuerzburg, Germany; Department of Neurology, Haga Teaching Hospital, The Hague, the Netherlands; Department of Neurosurgery, Haga Teaching Hospital, The Hague, the Netherlands; Department of Neurosurgery, Leiden University Medical Center, Leiden, the Netherlands; Department of Neurosurgery, Lausanne University Hospital (CHUV) and University of Lausanne (UNIL), Lausanne, Switzerland; Service of Neurology, Department of Clinical Neurosciences, Lausanne University Hospital (CHUV) and University of Lausanne (UNIL), Lausanne, Switzerland; Parkinson Institute of Milan, ASST G.Pini-CTO, Milan, Italy

**Keywords:** deep brain stimulation, local field potentials, beta power, contact prediction, Parkinson’s disease, directional lead

## Abstract

**Background:** The selection of optimal contacts for chronic deep brain stimulation (DBS) requires manual iterative testing of multiple stimulation configurations: the monopolar review. This requires time, highly trained personnel, and can cause patient discomfort. The use of neural biomarkers may help speed up this process.

**Objective:** This study aimed to validate the use of local field potentials (LFP) from a chronically implanted DBS neurostimulator to inform clinical selection of optimal stimulation contact-levels.

**Methods:** We retrospectively analysed bipolar LFP-recordings performed in patients with Parkinson’s disease OFF-medication and OFF-stimulation across three centres. For each contact-level chosen clinically, we ranked the recordings obtained by different channels according to the informative value of various beta-band (13-35Hz) power measures. We then developed two prediction algorithms: (i) a “decision-tree” method for direct, in-clinic use, and (ii) a “pattern based” method for offline validation. We finally compared these approaches to existing prediction algorithms.

**Results:** We included 68 subthalamic nuclei from the Netherlands (NL), 21 from Switzerland (CH), and 32 from Germany (DE). Recording channel rankings depended on the clinically chosen contact-level. When predicting the first two contact-levels, the online “decision tree” method achieved a predictive accuracy of 86.5% (NL), 86.7% (CH), and 75.0% (DE), respectively. The offline “pattern based” technique attained similar results. Both prediction techniques outperformed an existing algorithm and were robust in different clinical and recording conditions.

**Conclusion:** This study demonstrates that using these new methods, LFP-signals recorded in-clinic can accurately support the selection of stimulation contact-levels, showing potential to reduce DBS programming time.

## 1. Introduction

Deep brain stimulation (DBS) of the subthalamic nucleus (STN) is an effective treatment for Parkinson’s disease (PD) [1, 2]. Optimisation of stimulation parameters is critical to maximise therapy efficacy. Identification of the optimal stimulation contact-level is challenging and time-consuming, requiring long iterative manual testing of multiple combinations (monopolar review, MPR), and several follow-up visits [3]. Novel neurostimulator devices are now capable of recording local field potentials (LFP) from the chronically implanted DBS electrodes, which may serve as a neural biomarker to guide clinical programming [4–7]. LFP are generated by the integrated (weighted) sum of all synaptic potential changes [8]. These signals can provide direct insight into the functioning of basal ganglia circuits [8, 9]. Subthalamic LFP activity within the beta-frequency band (13-35 Hz) correlates with akinetic-rigid symptoms in PD and can be modulated by therapies (e.g., levodopa and STN-DBS) [10]. Suppression of power in the beta-frequency band typically corresponds with motor improvement in PD patients.

To date, various studies have explored LFP-based DBS contact-level selection techniques, mostly relying on neural features in the beta-band [11]. The maximum value of the beta-frequency power in a patient-specific range is mostly used as a measure to guide contact-level selection [12–19]. However, the clinical application of these techniques is still restricted due to limited validation, uncertainties regarding the reliability of the online visual inspection approach, or the need for complex methods and offline analysis. The most informative spectral feature of LFP for determining optimal stimulation therefore remains unclear.

Importantly, LFP activity is recorded bipolarly, between pairs of contact-levels. Identifying one individual monopolar stimulation level from these bipolar recordings presents a complex challenge. It has been suggested that either the contact-level in between the bipolar recording pair (e.g. contact-level 2 if channel (contact-level pair) 1-3 shows the highest maximum power) or one of the two recording contact-levels (e.g. contact-levels 1 or 3 if channel 1-3 shows the highest maximum power) could be the optimal stimulation location(s) [18, 19]. Studies addressing this issue using custom algorithms show a median predictive accuracy of 45% (range 25-71%) on selected datasets [20–26]. However, these algorithms often focus on predicting an optimal horizontal direction for stimulation [20–24] with directional leads, rather than an optimal vertical contact-level. Furthermore, they do not compare different predictors or techniques to derive monopolar predictions from bipolar recordings [20–25].

To better support initial DBS programming in PD, we explored the predictive efficacy of various beta-frequency band features using novel prediction techniques. We compared these novel techniques, which translate bipolar contact-level recordings to predictions for the optimal monopolar stimulation contact-level, with existing published algorithms.

## 2. Methods

### 2.1 Study design

This study concerns a retrospective, international multi-centre study across three DBS centres: Haga Teaching Hospital/Leiden University Medical Centre (The Netherlands (NL)); Lausanne University Hospital, (Switzerland (CH)); University Hospital of Würzburg (Germany (DE)). In the three participating centres, PD patients implanted in the subthalamic nucleus (STN) with directional Sensight™ leads in combination with the Percept PC® neurostimulator (Medtronic, Minneapolis) were screened for inclusion. In all three study centres ethical approval was obtained from local ethics committees.

### 2.2 Data acquisition

#### 2.2.1 LFP measurements

Contact-level bipolar LFP recordings conducted in the OFF-medication state (i.e. after overnight suspension of all dopaminergic drugs) with the *BrainSense™ Survey* (i.e. OFF-stimulation) were used. This research focussed on contact-level recordings (rings 0 and 3 and virtual rings 1abc and 2abc) of the directional Sensight™ lead, and did not evaluate individual segments (1a,-b,-c or 2a,-b,-c). All “NL” and “CH” recordings were performed within the first two weeks after lead implantation without previous DBS therapy, whereas “DE” measurements were mostly collected after a longer follow-up with a stimulation washout period of approximately 3 hours prior to MPR.

#### 2.2.2 Clinical contact-level choice

The contact-level chosen for chronic stimulation by the clinician after MPR served as a reference for all the LFP-based predictions. MPR was performed according to a standardised protocol between 8 days and 3 months after lead implantation (**Monopolar Review Protocol, Supplementary file-1**). For all centres, we additionally compared the level of the electrodes used for chronic stimulation at 1 year after surgery. In the case of interleaved programming at follow-up, we considered the contact with the highest stimulation amplitude, or, in the case of an equal contribution by both contacts, the contact with the smallest distance to the contact chosen at MPR. If directional stimulation was applied at follow-up the contact-level containing the active segment was considered.

### 2.3 Data analysis

Matlab (version R2022b, MathWorks^®^) was used for all data pre-processing and analyses.

#### 2.3.1 Data pre-processing and feature extraction

Available *Brainsense™ Survey* measurements were visually inspected for potential artefacts in the frequency and time domain. For the analyses, the frequency and power data derived from the time-to-frequency conversion embedded in the Percept PC® neurostimulator were used. Feature power values were thereafter determined by means of a custom Matlab script (<u>to_be_replaced_with_doi</u>). All analyses were performed for the left and right hemispheres separately. Stimulation contact-levels for both hemispheres were numbered 0, 1, 2 and 3 (ventral, ventromedial, dorsomedial, dorsal). When using the *Brainsense™ Survey* measurement bipolar recordings are conducted for the following six channels (contact-level pairs): 0-1; 0-2; 0-3; 1-2; 1-3; 2-3.

From all recording channels, four features of the beta-frequency band (13-35Hz) were extracted. The first is the maximum beta power value (“Max”), commonly used in clinical practice, as it is easily readable from the clinician programmer’s screen. As this value can be affected by varying levels of background noise across contacts, we also evaluated this feature after removal of the 1/f component (“Max_flat”) using the FOOF algorithm for a frequency range of 1 to 100Hz using the standard model settings (FOOOF v1.0.0 combined with the MATLAB wrapper v1.0.0) [27]. The third feature was the area under the curve over the whole beta-frequency band (“AUC”), which may be more representative or stable than a simple discrete value but requires offline analysis. This feature was additionally evaluated after removing the aperiodic 1/f component (“AUC_flat”) (**Figure 1**). For the main analyses, only *Brainsense™ Survey* recordings with a detectable beta-peak were included. The presence of the beta-peak was determined using a threshold for the “AUC_flat” value. In secondary analyses, the impact of limited beta-activity on the predictive accuracy of the proposed techniques was further assessed. Furthermore, in the subset of “NL” patients we evaluated the predictive accuracy of the techniques when individually focussing on the low-beta (13-20Hz) or high-beta (21-35Hz) peak activity (“Max” feature).

**Figure 1.**
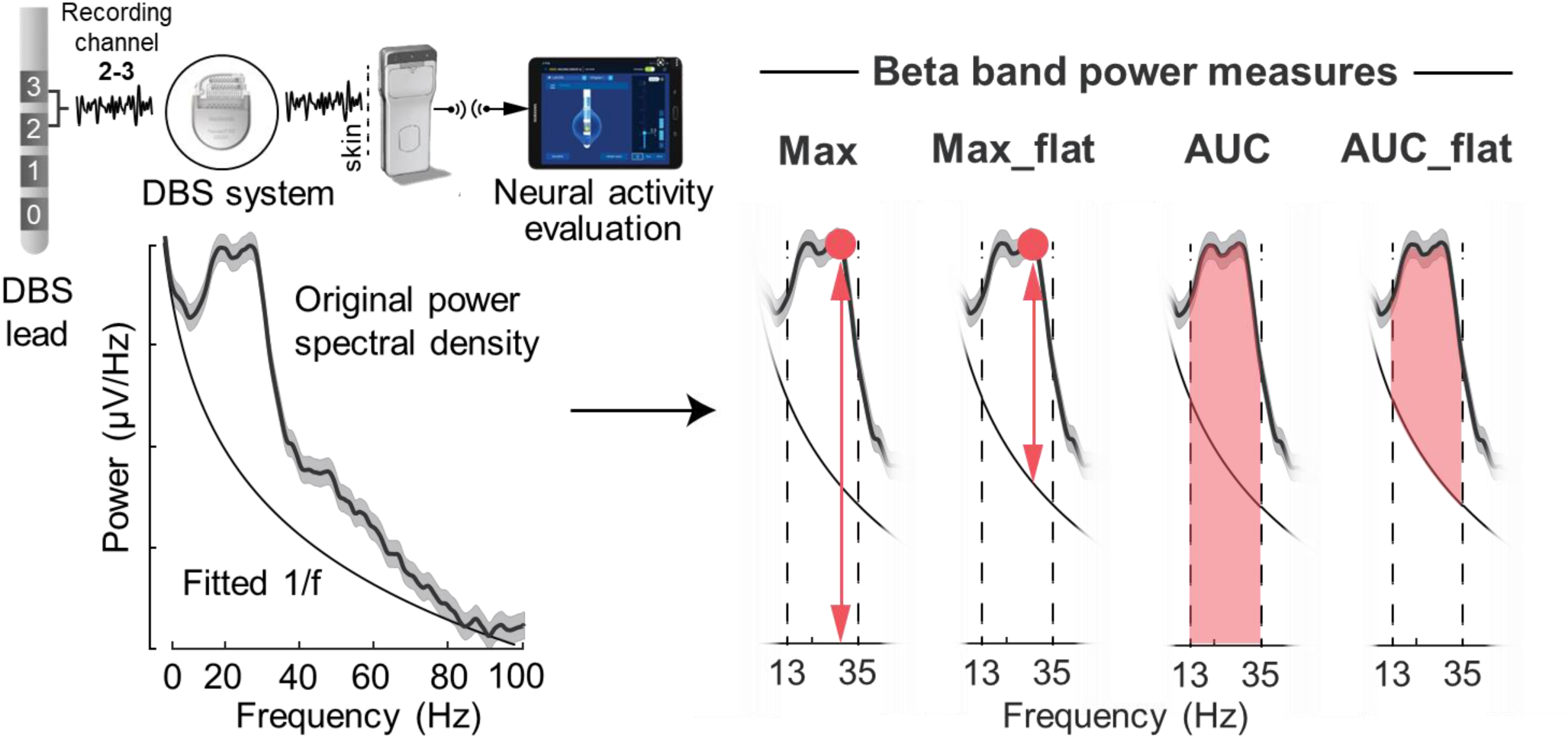
Original spectrum and FOOOF-algorithm aperiodic fit demonstrating the extraction of four beta-band power measures: the maximum power (Max); the maximum flattened (i.e. after removal of the aperiodic signal component) power (Max_flat); the area under the curve (AUC); and the flattened area under the curve (AUC_flat).

#### 2.3.2 From clinically-chosen contact-level to recording channel ranking

We aimed to determine the relation between bipolar recordings and clinical choice. To this end, for each clinically chosen contact-level across all hemispheres, we ranked all LFP recording channels based on the amplitude of each of the four aforementioned features individually (**Figure 4 and Supplementary figure 1, Supplementary file-2**).

**Figure 2.**
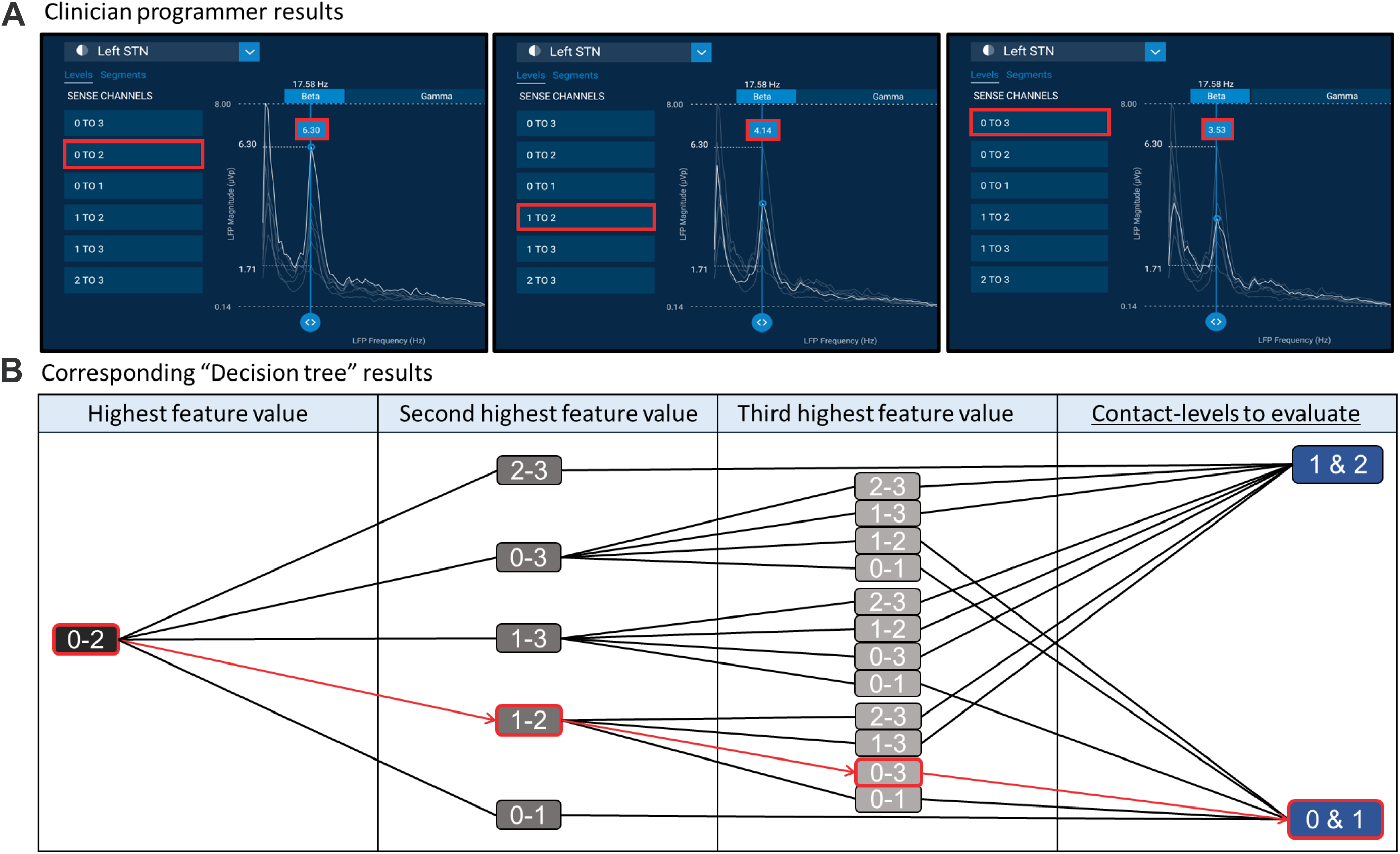
Example of the “selection decision tree” online ranking technique which uses the two or three recording channels with the highest beta feature value to select the two most likely best contact-levels. **A)** For this method the clinician performs a *BrainSense Survey* for the contact-levels using the clinician programmer. These contact-level recordings can thereafter be used on-screen to visually select the channel (contact pair) with the highest beta peak (i.e. channel “0 to 2” in this example). **B)**This information is used to choose the appropriate “selection decision tree”, i.e. the one which starts with the selected channel. Hereafter, the channel with the second highest beta peak (here channel “1 to 2”) is selected in the first branch of the decision tree. Finally, the channel with the third highest beta peak (here channel “0 to 3”) is selected in the second and final branch. The final block highlights the most promising contact-levels, identified here as levels 0 and 1. The large power between contact pairs 0-2 and 1-2 (highest and second-highest) could be due to either high power on contact _7_ 2 (and low on 0 and 1), or high power on contacts 0 and 1 (and low on 2). Because the third highest power is on contact pairs 0-3, this indicates that the large power is on 0 (and thus not on 2), only leaving out 0 and 1. These conclusions are derived using the “selection decision tree” technique only.

**Figure 3.**
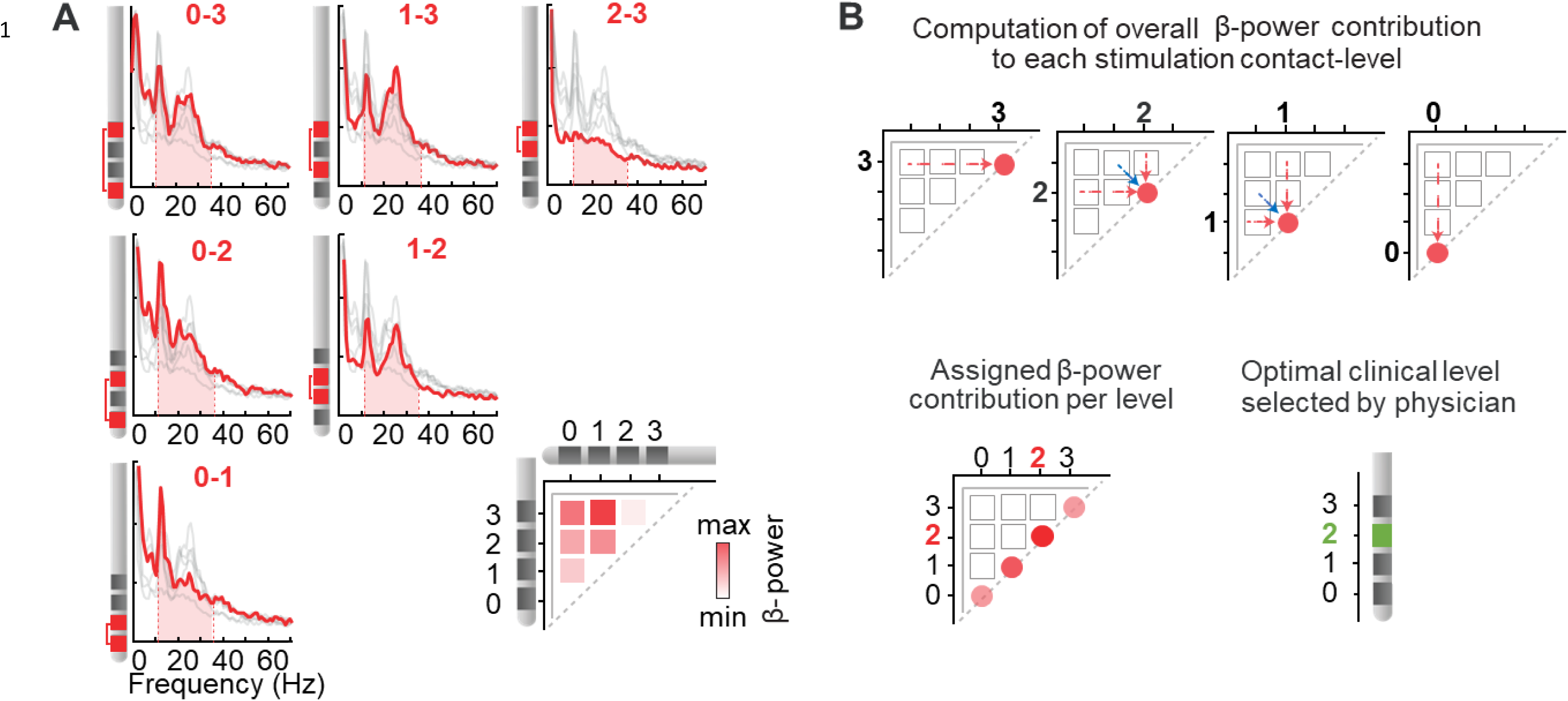
Example of the “pattern based” ranking technique which computes a single feature value per eligible contact-level to use for the selection of the optimal stimulation level. **A)** The feature value (example in figure = AUC) is determined for all recording channels (contact pairs) on one lead. This information is thereafter summarised in a single image where the intersection of contact-levels is represented on two axes using a heatmap with colour intensity corresponding to the feature value. (In this example maximum AUC power was at contact pairs 1-3 and minimum at contact pairs 2-3). **B)** The information in the summary image is thereafter converted using the “pattern based” method to provide a single “beta-power contribution” value for each of the four eligible contact-levels. This conversion is performed by: **1)** averaging all feature values for channels including a certain level and assigning this value to the respective contact-level (e.g. for level 0 feature values of: channel 0-1, channel 0-2 and channel 0-3 – red arrows - are averaged to obtain a single “beta-power contribution” value for level 0), and **2)** in addition, for contact levels 1 and 2, the feature power of the contact pairs adjacent to these levels (e.g. for level 1: channel 0-2, blue arrows) is compared to the values obtained in step 1) and the highest value between the two is assigned to these contact levels.This result is stored in a single “beta-power contribution” value per eligible contact-level. The level with the highest assigned value is then considered as the predicted optimal level (Here: level 2) . This prediction can then be compared to the optimal clinical level selected by the physician (green in the figure) to 9 determine the predictive accuracy of the “pattern based” technique.

**Figure 4.**
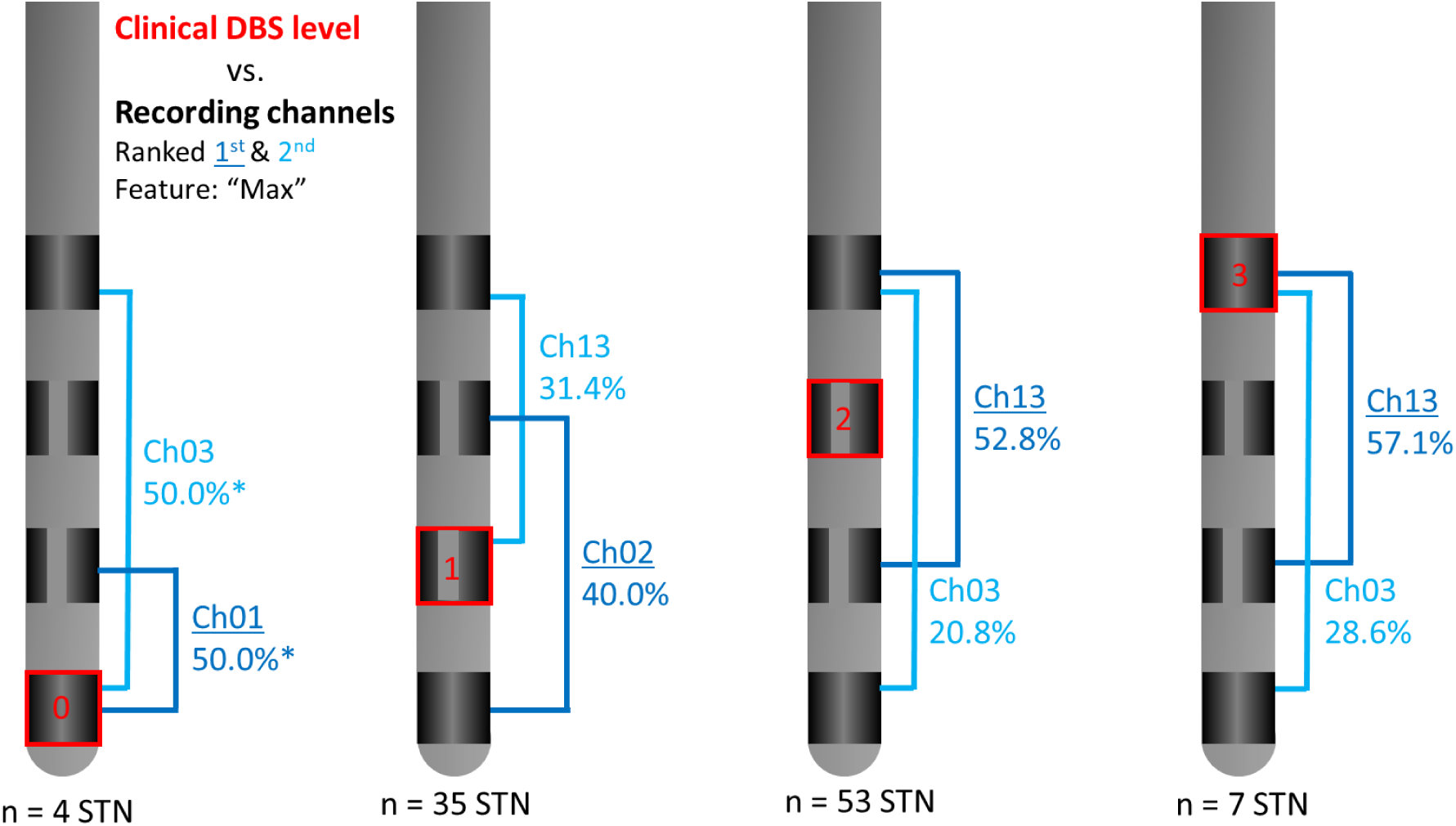
First- (dark blue) and second-ranked (light blue) recording channel (Ch) per clinically chosen deep brain stimulation (DBS) contact-level at monopolar review (in red) for the maximal power beta-frequency band feature (“Max”). This shows how often the channel was ranked first based on this feature across the entire population of subthalamic nuclei (STN) as a percentage per 1^st^ and 2^nd^ ranked channel. Results shown are for all included leads with clear beta activity. (*) In case of an equal percentage (i.e. channels are ranked first with equal frequency) the channel which was ranked second most often, of these two options, was indicated as ranked 1^st^.

#### 2.3.3 From recordings to contact-level prediction

To test the possibility of predicting the clinically-chosen contact-level directly from LFP recordings, two custom ranking methods were developed and evaluated, together with an existing ranking model (DETEC-algorithm [20]). All “NL” recordings were used to design the ranking methods (training set). External validation was performed using the “CH” and “DE” datasets (validation sets).

#### 2.3.3a. Ranking approach for online contact selection (“decision tree” method)

A ranking method based on a decision tree set of hierarchical models was developed as a tool to be used for online, in-clinic applications. The applied method is based on the theory that the feature value of the recording channels represents a difference in activity between two points along the lead. Based on this principle the decision tree is sequentially navigated using the channels with the first, second, and, when applicable, third highest feature values. The additional third step in the tree applies mostly when the second highest feature value leads to a channel with a large intercontact distance. By doing so the method accounts for differences in intercontact distance **(Intercontact Distance, Supplementary file-1**).

Two types of decision trees were included, a “selection tree” and an ”elimination tree”, that can be applied consecutively. The first decision tree (“selection tree”) uses the three bipolar recordings with the highest beta-peak (“Max” feature) to select the two best stimulation contact-levels (**Figure 2** and **Supplementary figure 1, Supplementary file-1**). The second decision tree (“elimination tree”) then discards the least promising stimulation levels by looking at the bipolar recordings with the lowest beta-peak (**Supplementary figure 2, Supplementary file-1**). We tested whether the use of the elimination tree in addition to the selection tree increased the accuracy.

This online selection technique was additionally tested for the three remaining features (AUC, Max_flat and AUC_flat). However, results relying on the “Max” feature are considered of greatest value as this method is most viable in an online clinical setting.

#### 2.3.3b Ranking approach for offline contact selection (“pattern based” method)

We developed a non-iterative method that maps the distribution of beta features across all bipolar recording pairs and provides an estimate of relevance for all stimulation contact candidates (<u>to_be_replaced_with_doi</u>). Albeit more computationally expensive, this “pattern based” process allows offline confirmation of the choice derived from the online decision-tree approach. Contributions to each stimulation contact may come from bipolar combinations that include the contact, as well as from pairs that surround the contact, our approach inherently accounts for both. Additionally, this method partially compensates for variations in intercontact distance by averaging across all distances and subsequently comparing results between contact-levels that share the same (average) distance **(Intercontact Distance, Supplementary file-1**).

In this “pattern based” method, beta features of all channels are first mapped onto a heatmap to represent the spatial distribution of the beta feature and the relative power that each bipolar recording channel conveys. This heatmap can then be used to derive the overall contribution of beta power to each contact-level by combining two methods. Method 1: averages the feature value across all channels which include the contact (e.g. for level 0: channel 0-1, channel 0-2 and channel 0-3); method 2: evaluates the feature value of the channels surrounding level 1 and 2 (e.g. for level 1: channel 0-2). The maximum value from both methods is kept, and the contact-level with the highest feature value is selected as the optimal DBS contact-level (**Figure 3**).

#### 2.3.3c Ranking approach for offline contact selection based on DETEC algorithm (Strelow et al. (2022)) [20]

We aimed to compare the results of both the online and offline methods to existing algorithms. As we only had access to level-based *Brainsense™ Survey* recordings, the only published algorithm that could be included was the DETEC-algorithm (**Equation 1**), which is an offline algorithm [20].

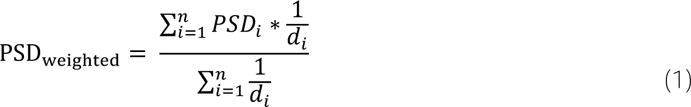

*In Equation(1), PSDi is the power spectral density from bipolar recording ί of the n bipolar recordings involving the investigated contact-level. dί is the distance between the centre of the investigated contact-level and its bipolar recording partner for each ίi-th bipolar recording (mm)*.

### 2.4 Model evaluation

To assess each ranking model’s performance, we calculated how often the clinically chosen stimulation level matched the 1^st^ ranked contact-level, or the 1^st^ or 2^nd^ ranked level (without any priority/weight), based on LFP. Including both the 1^st^ and 2^nd^ ranked levels allowed fair comparison between the “decision tree” method, which often selects two optimal contact-levels, and the other models, where a single optimal contact-level can be selected.

#### 2.4.1 Performance in case of limited beta power

Ranking models were additionally evaluated for sub-groups based on their amount of AUC beta activity without 1/f, (“AUC_flat” feature). Visual inspection defined that below a threshold of 0.6µV, the detection of a beta-peak was unclear. We thus clustered hemispheres into the following three groups: (i) “clear beta”: at least one channel with “AUC_flat” ≥ 0.6 µV. (ii) “little beta”: one or more channels with “AUC_flat” between 0.0 and 0.6 µV. (iii) “background signal only”: all channels with “AUC_flat” ≤ 0.0 µV. Even in cases with a flattened power (“little beta” or “background signal only”), we could still extract an order from the global beta (i.e., background activity in the beta-band) (**Supplementary figure 3, Supplementary file-1**). Therefore, ranking using the “AUC” feature is still possible in these cases, providing similar results as visual ranking of background activity in clinical practice.

#### 2.4.2 Performance in case of “Stun effect”

For the “NL” dataset, ranking models were evaluated for a subgroup of hemispheres with a stun effect at the time of MPR. A stun effect was considered when the symptoms were too mild to allow a reliable clinical evaluation at MPR (MDS-UPDRS-III OFF-medication score for bradykinesia, rigidity and tremor equal to 0 or 1).

#### 2.4.3 Effect of medication

The predictive accuracy of the algorithms was also assessed for a subset of LFP recordings (“CH” ON) where patients were ON-medication (i.e. practically defined ON).

### 2.5 Statistical analysis

Differences in population characteristics were evaluated using a Kruskal-Wallis test for numerical data or a Chi-Square test in case of categorical data. A Chi-square test was used to investigate significant differences in performance based on the subgroups described in 2.4.1 – 2.4.3.In all statistical evaluations, a two-sided p-value smaller than 0.05 was considered significant.

## 3. Results

### 3.1 Subject characteristics

Across the three centres, a *BrainSense™ Survey* measurement (after overnight suspension of all dopaminergic drugs) was available for 62 patients (121 STN) (**Table 1**). Additionally, an extra set of patients (“CH”, N=18, 35 STN) with recordings in ON-medication were included for a sub-analysis (**Supplementary table 5, Supplementary file-2**).

**Table 1.**
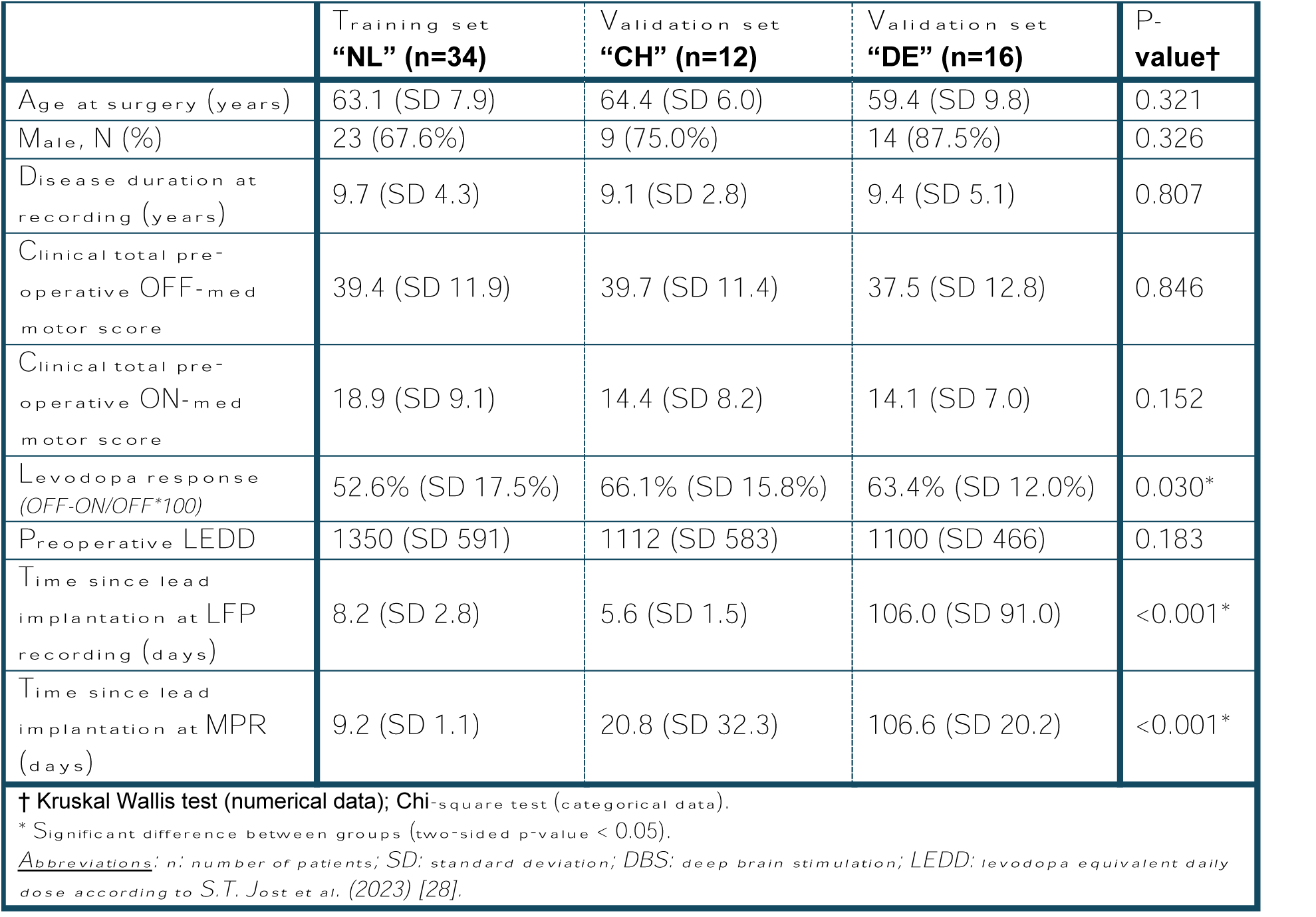
Patient characteristics.

|  | **Training set**<br>**“NL” (n=34)** | **Validation set**<br>**“CH” (n=12)** | **Validation set**<br>**“DE” (n=16)** | **P-**<br>**value†** |
| --- | --- | --- | --- | --- |
| Age at surgery (years) | 63.1 (SD 7.9) | 64.4 (SD 6.0) | 59.4 (SD 9.8) | 0.321 |
| Male, N (%) | 23 (67.6%) | 9 (75.0%) | 14 (87.5%) | 0.326 |
| Disease duration at recording (years) | 9.7 (SD 4.3) | 9.1 (SD 2.8) | 9.4 (SD 5.1) | 0.807 |
| Clinical total pre-operative OFF-med motor score | 39.4 (SD 11.9) | 39.7 (SD 11.4) | 37.5 (SD 12.8) | 0.846 |
| Clinical total pre-operative ON-med motor score | 18.9 (SD 9.1) | 14.4 (SD 8.2) | 14.1 (SD 7.0) | 0.152 |
| Levodopa response (OFF-ON/OFF*100) | 52.6% (SD 17.5%) | 66.1% (SD 15.8%) | 63.4% (SD 12.0%) | 0.030* |
| Preoperative LEDD | 1350 (SD 591) | 1112 (SD 583) | 1100 (SD 466) | 0.183 |
| Time since lead implantation at LFP recording (days) | 8.2 (SD 2.8) | 5.6 (SD 1.5) | 106.0 (SD 91.0) | <0.001* |
| Time since lead implantation at MPR (days) | 9.2 (SD 1.1) | 20.8 (SD 32.3) | 106.6 (SD 20.2) | <0.001* |
| † Kruskal Wallis test (numerical data); Chi-square test (categorical data).<br>* Significant difference between groups (two-sided p-value < 0.05).<br><u>Abbreviations:</u> n: number of patients; SD: standard deviation; DBS: deep brain stimulation; LEDD: levodopa equivalent daily dose according to S.T. Jost et al. (2023) [28]. |  |  |  |  |

The contact-level used for chronic stimulation at 1 year (or 6 months if 1 year was not reached) post-lead placement differed from the contact-level chosen during MPR in 6 out of 68 STN (9%) for the “NL” dataset, in 3 out of 21 STN (14%) for the “CH” dataset, and in 8 out of 32 STN (25%) for the “DE” dataset. No chronic contact choice was available for 20 STN (17%) in total (**Supplementary table 1, Supplementary file-2**). Since clinical choices showed minimal differences between MPR and chronic stimulation, the prediction methods were not evaluated using chronic contact choices as a reference.

### 3.2 Pre-processing

No ECG or other definite artefacts were detected upon visual inspection in any of the datasets (“NL”, “CH” and “DE”), therefore, no artefact extraction was applied.

Clear beta activity (based on “AUC_flat” threshold) was present in at least one recording channel for 80.5% of all leads (52/68 (76.5%) “NL”; 15/21 (71.5%) “CH”; 32/32 (100%) “DE”). The median value of AUC_flat” was 2.64 (range: -2.81 to 13.60).

### 3.3 From clinically-chosen contact-level to recording channel ranking

Rankings for the “Max” feature were similar to rankings obtained using the “AUC” feature. This similarity was also true for the “Max_flat” and “AUC_flat” rankings (**Figure 4 and Supplementary figure 1, Supplementary file-2**). Since the “Max” feature is most feasible for in-clinic use, we focus on this feature in the results.

The optimal contact chosen by clinicians across our three centres highlighted that contact-levels 1 and 2 were most selected (35 and 53 hemispheres respectively), whereas contact-levels 0 and 3 were only seldom employed (4 and 7 hemispheres, respectively). The ranking of the LFP channels indicated that for contact-levels 1 and 2, the sensing channels surrounding the stimulation levels (i.e. 0-2 and 1-3), most often showed the highest feature amplitude. The pattern in channel rankings for contact-levels 0 and 3 did not show such a similarity (**Figure 4**).

### 3.4 From recordings to contact-level prediction

We then evaluated the predictive accuracy of the 1^st^ and 2^nd^ ranked contact-levels defined by LFP, focusing on cases with clear beta activity. The “selection decision tree” method using the “Max” feature achieved 86.5% accuracy (“CH”: 86.7%; “DE”: 75.0%), increasing to 88.5% (“CH”: 93.3%; “DE”: 78.1%) when combining the selection and elimination trees (**Figure 5** and **Supplementary table 2, Supplementary file-2**). The “pattern based” method reached 84.6% accuracy (“CH”: 66.7%, “DE”: 71.9%) when considering both the 1^st^ and 2^nd^ ranked contact-levels.

**Figure 5.**
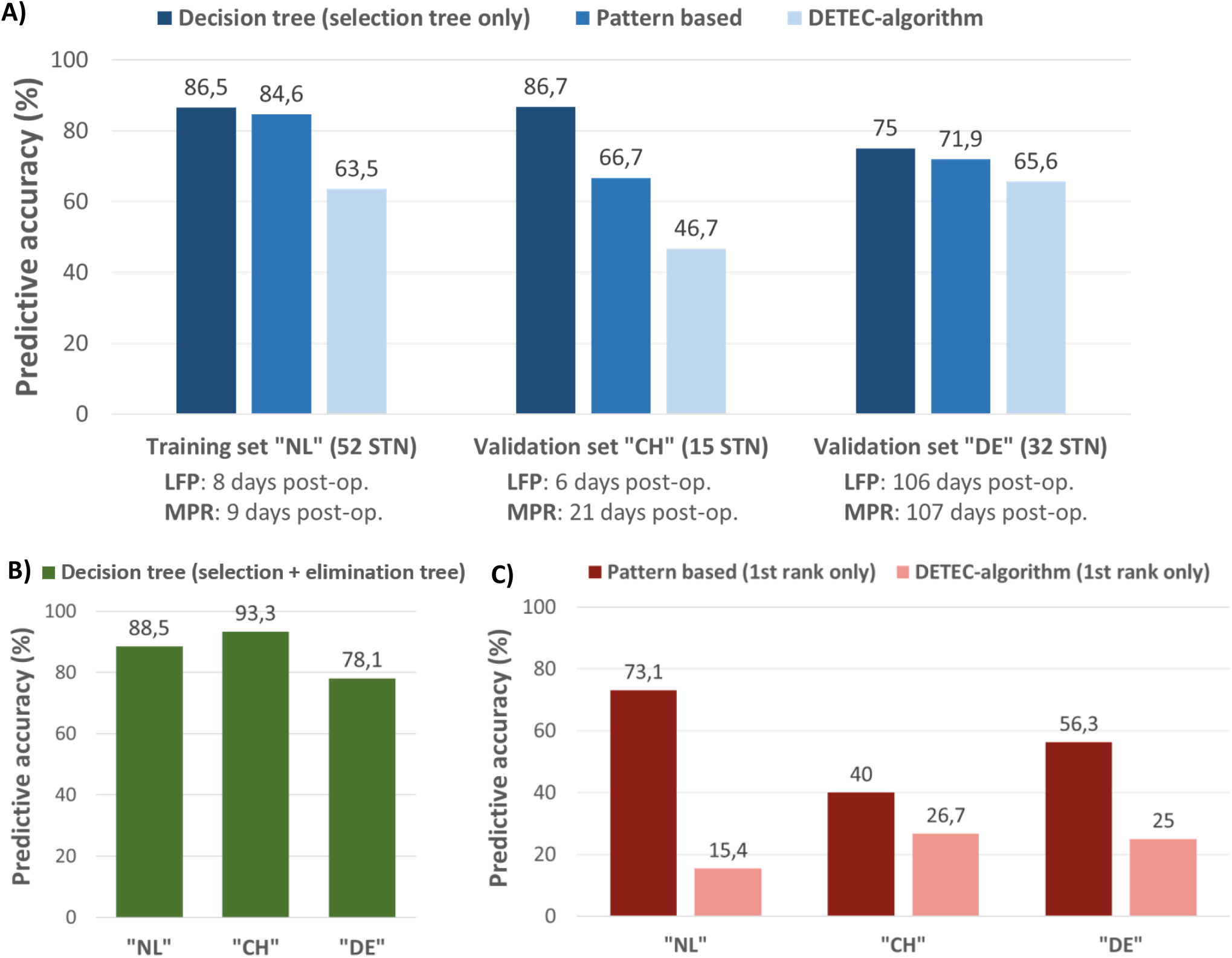
Comparison of algorithm performances when considering the results for the 1^st^ and 2^nd^ ranked contact-levels in recordings with clear beta activity obtained in OFF-medication. The time from the implant to LFP recordings (LFP: used for prediction) and the time from implant to the clinical contact selection (MPR: reference for prediction) varied between centres (see text). **A)** Predictive accuracy of the novel ‘decision tree – selection only’ and ‘pattern based’ prediction techniques for the ‘Max’ beta feature in comparison to the predictive accuracy of the existing DETEC-algorithm when considering both the 1^st^ and 2^nd^ ranked contact-levels. **B)** Predictive accuracy for the 1^st^ and 2^nd^ ranked contact-levels based on the online ‘decision tree’ method when applying the selection tree in combination with the elimination tree. **C)** Predictive accuracy of the offline “pattern based” method and DETEC-algorithms for the 1^st^ ranked contact-level alone.

Both the “pattern based” and DETEC algorithms performed better when combining 1^st^ and 2^nd^ ranked contact-levels instead of considering the 1^st^ ranked contact-level alone. However, the existing DETEC algorithm consistently showed lower predictive accuracies across all centres (**Figure 5** and **Supplementary table 3, Supplementary file-2**). Similar results were observed when only focussing on the low-beta or high-beta bands for the “decision tree” and “pattern based” methods (**Supplementary table 4, Supplementary file-2)**.

Analyses were repeated using the “Max_flat”, “AUC” and “AUC_flat” features for both the “decision tree” and “pattern based” methods. Changing the applied feature mostly resulted in minor differences in predictive accuracy. The “decision tree” method with both the selection and elimination tree in combination with the “AUC” feature achieved a slightly higher predictive accuracy in comparison to the “Max” feature for the “DE” dataset. The predictive accuracy across the “NL” and “CH” datasets was equal for both methods. When considering the “pattern based” method the “AUC_flat” feature slightly outperformed the use of the “Max” feature across the “NL” and “DE” datasets, and was equal for the “CH” dataset (**Supplementary tables 2, 3 and 5, Supplementary file-2**).

### 3.5 Subgroup comparison

#### 3.5.1. Performance in case of limited beta power

We then evaluated the predictive performance across hemispheres with little or no beta power above 1/f (**Table 2**). No significant differences were observed across any method or group. Across all data, containing all variations in beta-activity levels, the “decision tree” method, “pattern based” method and DETEC algorithm achieved predictive accuracies of 85.1%, 74.4% and 58.7%, respectively, when considering the 1^st^ and 2^nd^ ranked contact-levels.

**Table 2.** Frequency of 1^st^/2^nd^ ranked contact-levels corresponding to the clinically chosen stimulation level per ranking model for all patients from all three centres, subcategorised for clear or little beta above 1/f or background signal alone.

|  |  |  |  |
| --- | --- | --- | --- |
|  | **Decision tree<sup>†</sup>**<br>**1<sup>st</sup> / 2<sup>nd</sup> ranked** | **Pattern based**<br>**1<sup>st</sup> / 2<sup>nd</sup> ranked** | **DETEC algorithm**<br>**1<sup>st</sup> / 2<sup>nd</sup> ranked** |
| **Clear beta above 1/f**<br>("NL": 52 STN, "CH": 15 STN, "DE": 32 STN) | "Max": 82.8% (82) | "Max": 77.8% (77) | 61.6% (61) |
| **Little beta above 1/f**<br>("NL": 9 STN, "CH": 6 STN, "DE": 0 STN) | "AUC": 93.3% (14) | "AUC": 60.0% (9) | 46.7% (7) |
| **Background only**<br>("NL": 7 STN, "CH": 0 STN, "DE": 0 STN) | "AUC": 100.0% (7) | "AUC": 57.1% (4) | 42.9% (3) |
| **Chi-square\*** | $\chi^2 = 2.434$, $p = 0.296$ | $\chi^2 = 3.319$, $p = 0.190$ | $\chi^2 = 1.968$, $p = 0.374$ |
| <sup>†</sup> Results for selection decision tree only.<br>* Chi-square test was considered significant if two-sided p-value < 0.05<br>*Abbreviations: "Max": maximum power feature; "AUC": area under the curve feature* |  |  |  |

#### 3.5.2 Performance in case of “stun effect”

A sub-analysis was performed for all “NL” recordings without clinically detectable stun effect during MPR (**Table 3**). Of the 26 STN with a stun effect, 11 (42.3%) also showed limited LFP-based beta activity (i.e. little beta above 1/3 or background activity only).

**Table 3.** Frequency of 1^st^/2^nd^ ranked contact-levels corresponding to the clinically chosen stimulation level per ranking model for all “NL” patients, subcategorised as with/without stun effect.

|  |  |  |  |
| --- | --- | --- | --- |
|  | **Decision tree<sup>†</sup> –**<br>**“Max/AUC”**<br>**1<sup>st</sup> / 2<sup>nd</sup> ranked** | **Pattern based –**<br>**“Max/AUC”**<br>**1<sup>st</sup> / 2<sup>nd</sup> ranked** | **DETEC algorithm**<br>**1<sup>st</sup> / 2<sup>nd</sup> ranked** |
| **No stun**<br>(42 STN) | 83.3% (35) | 85.7% (36) | 66.7% (28) |
| **With stun**<br>(26 STN) | 96.2% (25) | 65.4% (17) | 46.2% (12) |
| **Chi-square\*** | $\chi^2 = 2.543$, p = 0.111 | $\chi^2 = 3.860$, p = 0.049 | $\chi^2 = 2.790$, p = 0.095 |
| † Results for selection decision tree only.<br>* Chi-square test was considered significant if two-sided p-value < 0.05<br>Abbreviations: Max: maximum power feature |  |  |  |

Patients with a stun effect had significantly lower predictive accuracy using the “pattern based” algorithm with the “Max” (if clear beta-activity) or “AUC” feature (if little beta-activity or background signals alone). The stun effect did not have a significant effect on the performance of the DETEC algorithm or “decision tree” method. No significant differences were found in patient characteristics, except for a lower pre-operative OFF-medication motor score (p = 0.029) in the stun effect group (**Supplementary table 6, Supplementary file-2**).

#### 3.5.3 Effect of medication

Predictive accuracy differences between ON- and OFF-medication states were evaluated on an additional ON-medication “CH” dataset (**Table 4**). There were no significant differences in predictive accuracy or patient characteristics between subgroups (**Supplementary tables 7-8, Supplementary file-2**).

**Table 4.** Frequency of 1^st^/2^nd^ ranked contact-levels corresponding to the clinically chosen stimulation level per ranking model for all “CH” patients, for the ON-versus OFF-medication state.

|  |  |  |  |
| --- | --- | --- | --- |
|  | **Decision tree† - “Max”**<br>**1<sup>st</sup> / 2<sup>nd</sup> ranked** | **Pattern based – “Max”**<br>**1<sup>st</sup> / 2<sup>nd</sup> ranked** | **DETEC algorithm**<br>**1<sup>st</sup> / 2<sup>nd</sup> ranked** |
| **OFF-medication**<br>(21 STN) | 85.7% (18) | 66.7% (14) | 47.6% (10) |
| **ON-medication**<br>(35 STN) | 71.4% (25) | 68.6% (24) | 51.4% (18) |
| **Chi-square\*** | $\chi^2 = 1.503$, p = 0.220 | $\chi^2 = 0.022$, p = 0.883 | $\chi^2 = 0.076$, p = 0.783 |
| † Results for selection decision tree only.<br>* Chi-square test was considered significant if two-sided p-value < 0.05<br>Abbreviations: Max: maximum power feature |  |  |  |

## 4. Discussion

### 4.1 Algorithm performance and comparison to existing approaches

We retrospectively evaluated LFP signals and their relation to clinically chosen stimulation contact-levels across three European centres and developed a novel algorithmic framework, including two complementary prediction algorithms, to identify optimal DBS contact-levels in over 60 PD patients.

The decision-tree approach using the “Max” feature showed high predictive accuracy (>85%) for the 1^st^ and 2^nd^ optimal contact-level predictions combined and showed robustness across validation sets. This new, online technique can thus help restrict the search field for optimal stimulation contacts to two contact-levels for the majority of patients, thereby showing the potential to reduce DBS programming time. Incorporating elimination trees improved results by 2 to 6 percental points, suggesting that this extra step might not be crucial and will only be reserved for selected cases.

The “pattern based” approach using the same feature correctly predicted the single optimal contact-level in 73.1% of the cases, and reached near 85% accuracy for combining 1^st^ and 2^nd^ optimal contact-level predictions. These results were, however, slightly less robust across validation sets compared to the online “decision tree” method.

Both approaches outperformed the existing DETEC algorithm for our datasets. However, we note that the predictive accuracy achieved by the DETEC-algorithm in the original research was higher than the predictive accuracy found on our data [20]. The results from the “DE” dataset resemble the original results reported for the DETEC-algorithm most, likely because both datasets were recorded over 3 months post-lead placement.

Other existing algorithms using contact-level *BrainSense™ Survey* recordings (e.g. [25]) were not tested here, as they are not publicly available to our knowledge, and, based on the available literature, their accuracy is surpassed by the DETEC algorithm [20, 25]. Other existing algorithms are based on segmented *BrainSense™ Survey* recordings, which were not available in this retrospective analysis [22–24, 29].

### 4.2 Relevance of beta features for prediction performance

In all our analyses, using the “Max” and “AUC” feature yielded similar results. This indicates that albeit a single maximal value ignores the underlying PSD shape and is more prone to noise corruption, the conveyed information is enough for programming purposes. Once the aperiodic signal component is removed, the channels ranked second changed to channels with a smaller pick-up area (e.g. for contact-level 2 channel 0-3 is ranked second using the “Max” feature, whereas channel 2-3 is ranked second using the “Max_flat” feature). These changes were similar for both the “Max” and “AUC” features. Furthermore, the ranking of the LFP channels indicated that for contact-levels 1 and 2, the sensing channels surrounding the stimulation levels (i.e. 0-2 and 1-3), most often showed the highest feature amplitude. The pattern in channel rankings for contact-levels 0 and 3 did not show such a similarity.

For the online “decision tree” method, the features without removal of 1/f outperformed the flattened features. This is very convenient, since visual inspection of a single point is easier and faster, especially in the context of clinical evaluations. However, the opposite was true for the offline “pattern based” method.

### 4.3 Robustness across different clinical and recording conditions

The presence of beta activity is often regarded as one of the limiting factors when considering beta power predictions. We thus clustered LFP recordings into subsets depending on their amount of beta power (clear beta-activity, little beta-activity, and background signal alone). While visually setting a threshold for minimal beta activity is inherently subjective, this approach aligns with clinical practice. Furthermore, no significant difference in performance was observed across the different levels of beta-activity for the proposed algorithms. Nonetheless, it is important to emphasize that the proposed methods depend on the presence of an electrophysiological biomarker, and that this limitation inherent to electrophysiology-based contact choice, restricts the applicability of these techniques across patients.

A second sub-analysis on the “NL” dataset alone showed that the performance of the “decision tree” method and DETEC-algorithm remained stable even in patients with a clinical stun effect during LFP recordings, whereas the performance decreased significantly for the “pattern based” method.

Across our three centres, the amount of AUC beta-activity without 1/f (“AUC_flat” feature) correlated with the time at which LFP recordings were performed. All the recordings with little beta activity or background signal alone were seen in the “NL” and “CH” datasets, for which LFP recordings were performed at an average of 8 and 6 days after implantation, respectively. The “DE” dataset, for which LFP recordings were performed at an average of 106 days after lead implantation, only contained LFP recordings with clear beta-activity.

In the “CH” ON-medication dataset, accuracy was similar with and without medication, opening up the possibility of avoiding overnight medication withdrawal in the future.

While both methods, especially the “decision tree”, were generally robust in different clinical and recording conditions, the “decision tree” method using the “Max” feature may be more reliable in the OFF-medication state and post-stun effect in individual patients. Adding the elimination tree could enhance prediction validity in selected cases, and further confirmation can be obtained using the “pattern based” method with the “AUC_flat” feature.

### 4.4 Limitations

The studied population comprises a large cohort across three different centres, but limitations in the generalisation of the results should be considered. In the “NL” dataset, none of the STN were stimulated through contact-level 0, and only four (5.9%) through contact-level 3. This reflects the preferences and accuracy of the lead placement, as during surgery it is attempted to place the middle of the lead (contact-levels one and two, capable of steering) within the STN. This positioning is thereafter additionally confirmed using post-operative anatomical validation. This leads to contact-levels one and two being placed most optimally, explaining why these contact-levels are chosen for clinical stimulation with the highest frequency. Similar clinical selection patterns were seen in the other two datasets (“DE” level-0: 11.8%, level-3: 11.8%; “CH” level-0: 0.0%, level-3: 0.0%). Additionally, although no artefacts were visually identified, there may have been artefacts overlapping with the neurophysiological signal may have been present. Furthermore, differences in impedances between recording channels were not considered. Similarly, variations in distances between recording levels were not directly corrected in the new techniques, unlike the DETEC-algorithm. However, these distance variations were indirectly addressed through averaging in the “pattern based” method and through specific branch designs in the “decision tree” method.

The developed methods were not assessed for identifying the optimal stimulation segment in directional contacts, as segmented *BrainSense™ Survey* recordings were unavailable for most patients. By contrast, other algorithms, such as the DETEC algorithm, often support optimal segment identification. Nonetheless, the choice of directionality forms a second step in identifying the optimal stimulation, and is not needed in many individuals at the time of MPR [30]. At this time our methods do not include directionality, but do enable a fast choice among the available contact-levels, without the need for longer, additional segmented *BrainSense™ Survey* recordings.

Another important point is that bipolar recordings reflect the difference in electrical activity between two recording points, making direct translation to a single location with the highest activity inherently complex. Crucially, our work represents one of several efforts to propose a viable method for translating bipolar recordings to monopolar contact-level selection. However, such translations will inherently involve some degree of estimation. The future availability of (pseudo)monopolar LFP recordings may enhance prediction accuracy by removing the need for bipolar-to-monopolar translation, although this shift may introduce new challenges for implanted systems. Despite these advancements, bipolar recordings will remain essential during therapeutic stimulation and are likely to continue playing a pivotal role in the development of future adaptive systems.

We evaluated beta-based features within the 13-35Hz range, which differs from the beta range shown by the clinician programmer (8-30Hz). Additionally, a distinction between low- and high-beta activity was not made for all analyses, as initial results showed no clear difference in predictive accuracy.

Furthermore, not all patients exhibit a clear separation between low- and high-beta peak, as many patients showed a single peak around 20-21Hz. In these cases separating low- and high-beta bands would artificially lead to two “Max” and “Max_flat” features, biasing the results. Moreover, several patients only showed either low- or high-beta peaks but not both. This further illustrates that the distinction between low- and high-beta can be very complex and that specifically developing an algorithm for the individual low- or high-beta band would complicate the clinical workflow and reduce the clinical applicability. In this study we focussed mainly on the full beta-band, aiming to develop a simple and robust method that can be readily implemented in clinical practice. However, not all patients show a distinct relation between beta suppression and motor improvement [31–33]. Moreover, features from other frequencies may aid in predicting optimal stimulation levels [13, 34–36].

After MPR, the contact-level for chronic stimulation is typically selected based on the lowest threshold for therapeutic benefit, while also considering the absence of side-effects. This implies that even if a contact has the lowest threshold for benefit, it might still be excluded due to the presence of side-effects—an aspect that may not be fully reflected by LFP measurements. In this retrospective study, we were unable to verify this, as some necessary data were unavailable.

Additionally, if a stun effect occurs during MPR, the clinical contact level might be chosen based on other considerations, such as anatomical factors, personal preferences of the physician. Selecting a contact based on these other alternative factors could potentially lead to a choice that is suboptimal for stimulation. To correct for this, we also considered the contact-level used for stimulation 6-12 months after implant and found few differences (in 9-13-≤25% of STN). Finally, even though the Percept PC^®^ sensing capabilities were programmatically not used to select the most optimal contact-level during clinical programming, we cannot guarantee that physicians were blinded to the results of the sensing due to the retrospective character of the study. However, the fact that across all STN 86% of the contacts selected for chronic stimulation remained unchanged after 6-12 months further confirms that the clinical contact-level choice during MPR was indeed clinically optimal.

### 4.5 Conclusions and future directions

This study introduces and validates both online and offline tools for supporting selection of optimal clinical contact-levels, potentially reducing the time required for the initial optimal contact-level selection. The presented novel approaches are robust under varying clinical conditions. The online decision tree is particularly practical for clinical use.

Future research should explore including directionality using the segmented *BrainSense™ Survey* recordings, and should consider incorporating features from other frequency bands, which may relate to certain PD symptoms and/or disease states [37]. In addition, integrating our methods with imaging data could provide information on the expected locations of side-effects. Including information on stimulation-induced beta-suppression could further enhance optimal contact-level selection [21, 23]. Future availability of monopolar LFP recordings might also improve predictions by eliminating the need for a bipolar to monopolar translation, although new challenges may arise in implanted systems.

While the proposed methods can enhance LFP-based contact-level selection and reduce testing time, especially in case of clinical complexity, clinical judgement and case-by-case considerations should always remain.

## CRediT roles

- MM: Conceptualisation; Methodology; Software; Validation; Formal analysis; Investigation; Data Curation; Writing - Original Draft; Visualization.
- SS: Software; Formal Analysis; Investigation; Writing - Review & Editing.
- IH: Formal Analysis; Investigation; Writing - Review & Editing.
- CV: Formal Analysis; Investigation; Writing - Review & Editing.
- CP: Software; Formal Analysis; Investigation; Writing - Review & Editing.
- SvdG / RZ / NAvdG / CFEH / JB / MCJ / JFB / PC: Writing - Review & Editing.
- IUI: Conceptualisation; Methodology; Writing - Review & Editing; Supervision.
- EMM: Conceptualisation; Methodology; Software; Formal Analysis; Investigation; Writing – Review & Editing; Visualisation; Supervision.
- MFC: Conceptualisation; Methodology; Formal Analysis; Investigation; Writing – Review & Editing; Visualisation; Supervision.

## Data Availability

The datasets used and analysed during the current study available from the corresponding author on reasonable request. The underlying code for this study is available in the Open Science Framework.

https://osf.io/fzn8d/?view_only=e2ea2e82f3664989b7e8a5d4b3c1cec2

## Acknowledgements

The authors are thankful to Prof.dr.ir. A.C. Schouten and his team for helpful insights.

## Data availability statement

The datasets used and analysed during the current study available from the corresponding author on reasonable request.

## Code Availability Statement

The underlying code for this study is available in the Open Science Framework and can be accessed via this link https://osf.io/fzn8d/?view_only=e2ea2e82f3664989b7e8a5d4b3c1cec2.

## References

[1] Ince NF, Gupte A, Wichmann T, Ashe J, Henry T, Bebler M, et al. Selection of optimal programming contacts based on local field potential recordings from subthalamic nucleus in patients with Parkinson’s disease. Neurosurgery. 2010;67(2):390–7.

[2] Pintér D, Járdaházi E, Balás I, Harmat M, Makó T, Juhász A, et al. Antiparkinsonian Drug Reduction After Directional Versus Omnidirectional Bilateral Subthalamic Deep Brain Stimulation. Neuromodulation. 2023;26(2):374–81.

[3] Volkmann J, Moro E, Pahwa R. Basic algorithms for the programming of deep brain stimulation in Parkinson’s disease. Mov Disord. 2006;21 Suppl 14:S284–9.

[4] Medtronic. PERCEPT PC-NEUROSTIMULATOR medtronic.eu20202020 [Available from: https://europe.medtronic.com/xd-en/healthcare-professionals/products/neurological/deep-brain-stimulation-systems/percept-pc.html.

[5] Isaias IU, Caffi L, Borellini L, Ampollini AM, Locatelli M, Pezzoli G, et al. Case report: Improvement of gait with adaptive deep brain stimulation in a patient with Parkinson’s disease. Front Bioeng Biotechnol. 2024;12:1428189.

[6] Caffi L, Romito LM, Palmisano C, Aloia V, Arlotti M, Rossi L, et al. Adaptive vs. Conventional Deep Brain Stimulation: One-Year Subthalamic Recordings and Clinical Monitoring in a Patient with Parkinson’s Disease. Bioengineering [Internet]. 2024; 11(10).

[7] Arlotti M, Palmisano C, Minafra B, Todisco M, Pacchetti C, Canessa A, et al. Monitoring subthalamic oscillations for 24 hours in a freely moving Parkinson’s disease patient. Movement Disorders. 2019;34(5):757–9.

[8] Lashgari R, Li X, Chen Y, Kremkow J, Bereshpolova Y, Swadlow HA, et al. Response properties of local field potentials and neighboring single neurons in awake primary visual cortex. J Neurosci. 2012;32(33):11396–413.

[9] Einevoll GT, Kayser C, Logothetis NK, Panzeri S. Modelling and analysis of local field potentials for studying the function of cortical circuits. Nat Rev Neurosci. 2013;14(11):770–85.

[10] Brittain J-S, Brown P. Oscillations and the basal ganglia: Motor control and beyond. NeuroImage. 2014;85:637–47.

[11] van Wijk BCM, de Bie RMA, Beudel M. A systematic review of local field potential physiomarkers in Parkinson’s disease: from clinical correlations to adaptive deep brain stimulation algorithms. J Neurol. 2023;270(2):1162–77.

[12] Xu SS, Lee WL, Perera T, Sinclair NC, Bulluss KJ, McDermott HJ, et al. Can brain signals and anatomy refine contact choice for deep brain stimulation in Parkinson’s disease? J Neurol Neurosurg Psychiatry. 2022.

[13] Xu SS, Sinclair NC, Bulluss KJ, Perera T, Lee WL, McDermott HJ, et al. Towards guided and automated programming of subthalamic area stimulation in Parkinson’s disease. Brain Commun. 2022;4(1):fcac003.

[14] Sinclair NC, McDermott HJ, Lee WL, Xu SS, Acevedo N, Begg A, et al. Electrically evoked and spontaneous neural activity in the subthalamic nucleus under general anesthesia. J Neurosurg. 2021:1–10.

[15] Milosevic L, Scherer M, Cebi I, Guggenberger R, Machetanz K, Naros G, et al. Online Mapping With the Deep Brain Stimulation Lead: A Novel Targeting Tool in Parkinson’s Disease. Mov Disord. 2020;35(9):1574–86.

[16] Tamir I, Wang D, Chen W, Ostrem JL, Starr PA, de Hemptinne C. Eight cylindrical contact lead recordings in the subthalamic region localize beta oscillations source to the dorsal STN. Neurobiol Dis. 2020;146:105090.

[17] Tinkhauser G, Pogosyan A, Debove I, Nowacki A, Shah SA, Seidel K, et al. Directional local field potentials: A tool to optimize deep brain stimulation. Mov Disord. 2018;33(1):159–64.

[18] Kochanski RB, Shils J, Verhagen Metman L, Pal G, Sani S. Analysis of Movement-Related Beta Oscillations in the Off-Medication State During Subthalamic Nucleus Deep Brain Stimulation Surgery. J Clin Neurophysiol. 2019;36(1):67–73.

[19] Thenaisie Y, Palmisano C, Canessa A, Keulen BJ, Capetian P, Castro Jimenez M, et al. Towards adaptive deep brain stimulation: clinical and technical notes on a novel commercial device for chronic brain sensing. Journal of neural engineering. 2021;13.

[20] Strelow JN, Dembek TA, Baldermann JC, Andrade P, Jergas H, Visser-Vandewalle V, et al. Local Field Potential-Guided Contact Selection Using Chronically Implanted Sensing Devices for Deep Brain Stimulation in Parkinson’s Disease. Brain Sci. 2022;12(12).

[21] Strelow JN, Dembek TA, Baldermann JC, Andrade P, Fink GR, Visser-Vandewalle V, et al. Low beta-band suppression as a tool for DBS contact selection for akinetic-rigid symptoms in Parkinson’s disease. Parkinsonism Relat Disord. 2023;112:105478.

[22] Binder T, Lange F, Pozzi N, Musacchio T, Daniels C, Odorfer T, et al. Feasibility of local field potential-guided programming for deep brain stimulation in Parkinson’s disease: A comparison with clinical and neuro-imaging guided approaches in a randomized, controlled pilot trial. Brain Stimulation. 2023;16(5):1243–51.

[23] Busch JL, Kaplan J, Bahners BH, Roediger J, Faust K, Schneider GH, et al. Local Field Potentials Predict Motor Performance in Deep Brain Stimulation for Parkinson’s Disease. Movement Disorders. 2023.

[24] di Biase L, Piano C, Bove F, Ricci L, Caminiti ML, Stefani A, et al. Intraoperative Local Field Potential Beta Power and Three-Dimensional Neuroimaging Mapping Predict Long-Term Clinical Response to Deep Brain Stimulation in Parkinson Disease: A Retrospective Study. Neuromodulation. 2023.

[25] Lewis S, Radcliffe E, Ojemann S, Kramer DR, Hirt L, Case M, et al. Pilot Study to Investigate the Use of In-Clinic Sensing to Identify Optimal Stimulation Parameters for Deep Brain Stimulation Therapy in Parkinson’s Disease. Neuromodulation. 2023.

[26] Dong W, Qiu C, Chang L, Sun J, Yan J, Luo B, et al. The guiding effect of local field potential during deep brain stimulation surgery for programming in Parkinson’s disease patients. CNS Neuroscience and Therapeutics. 2023.

[27] Donoghue T, Haller M, Peterson EJ, Varma P, Sebastian P, Gao R, et al. Parameterizing neural power spectra into periodic and aperiodic components.

[28] Jost ST, Kaldenbach M-A, Antonini A, Martinez-Martin P, Timmermann L, Odin P, et al. Levodopa Dose Equivalency in Parkinson’s Disease: Updated Systematic Review and Proposals. Movement Disorders. 2023;38(7):1236–52.

[29] Niso G, Tadel F, Bock E, Cousineau M, Santos A, Baillet S. Brainstorm Pipeline Analysis of Resting-State Data From the Open MEG Archive. Front Neurosci. 2019;13:284.

[30] Zitman FMP, Janssen A, van der Gaag NA, Hoffmann CFE, Zutt R, Contarino MF. The actual use of directional steering and shorter pulse width in selected patients undergoing deep brain stimulation. Parkinsonism Relat Disord. 2021;93:58–61.

[31] Ozturk M, Abosch A, Francis D, Wu J, Jimenez-Shahed J, Ince NF. Distinct subthalamic coupling in the ON state describes motor performance in Parkinson’s disease. Mov Disord. 2020;35(1):91–100.

[32] Kühn AA, Tsui A, Aziz T, Ray N, Brücke C, Kupsch A, et al. Pathological synchronisation in the subthalamic nucleus of patients with Parkinson’s disease relates to both bradykinesia and rigidity. Exp Neurol. 2009;215(2):380–7.

[33] Hill ME, Johnson LA, Wang J, Escobar Sanabria D, Patriat R, Cooper SE, et al. Paradoxical Modulation of STN β-Band Activity with Medication Compared to Deep Brain Stimulation. Mov Disord. 2024;39(1):192–7.

[34] Nguyen TAK, Schüpbach M, Mercanzini A, Dransart A, Pollo C. Directional Local Field Potentials in the Subthalamic Nucleus During Deep Brain Implantation of Parkinson’s Disease Patients. Front Hum Neurosci. 2020;14:521282.

[35] Connolly AT, Kaemmerer WF, Dani S, Stanslaski SR, Panken E, Johnson MD, et al., editors. Guiding deep brain stimulation contact selection using local field potentials sensed by a chronically implanted device in Parkinson’s disease patients. 2015 7th International IEEE/EMBS Conference on Neural Engineering (NER); 2015: IEEE.

[36] Shah A, Nguyen TK, Peterman K, Khawaldeh S, Debove I, Shah SA, et al. Combining Multimodal Biomarkers to Guide Deep Brain Stimulation Programming in Parkinson Disease. Neuromodulation. 2023;26(2):320–32.

[37] Yin Z, Zhu G, Zhao B, Bai Y, Jiang Y, Neumann WJ, et al. Local field potentials in Parkinson’s disease: A frequency-based review. Neurobiol Dis. 2021;155:105372.

