## Supplementary file 1 for "Online prediction of optimal deep brain stimulation contacts from local field potentials in chronically-implanted patients with Parkinson’s disease"

### Monopolar Review Protocol

The monopolar review (MPR) protocol was used for clinical contact-level selection in deep brain stimulation (DBS). This evaluation was conducted in the OFF-medication state (i.e. after overnight suspension of all dopaminergic drugs) to assess the effects and side-effects of stimulation for each individual contact-level. The stimulation current was gradually increased during MPR to identify the optimal contact-level.

**Procedure**

1. Baseline Assessment
    - Before stimulation, bradykinesia, rigidity, and tremor were scored using the Unified Parkinson's Disease Rating Scale Part III (UPDRS-III) as a baseline measure.
2. Incremental Stimulation
    - Stimulation was applied to a single contact-level at a time.
    - The stimulation current was progressively increased until a predefined maximum amplitude or until interfering side-effects were observed.
3. Evaluation during Stimulation
    - The following were recorded during progressive stimulation:
        - UPDRS-III scores for bradykinesia, rigidity, and tremor.
        - Any side effects or increases in side-effect severity reported by the patient.
4. Therapeutic Window Identification
    - After evaluating all contact-levels within a single hemisphere according to the above mentioned steps, the therapeutic window was determined for each contact-level.
        - The therapeutic window is defined as the range between:
            - The stimulation current required to achieve beneficial effects.
            - The stimulation current at which adverse effects occur.
    - Using this information the contact-level was then selected for chronic stimulation.
5. Repeat for Opposite Hemisphere
    - The process was repeated for the opposite hemisphere.
    - MPR was often initiated in the hemisphere with the most severe symptoms.

### Intercontact Distance

For the "pattern based" technique all bipolar recordings of a respective contact are averaged to calculate it's feature power. For contact 1 and 2 we then compare this averaged value with that obtained by recording channel 0-2 or 1-3, respectively. The highest value (average or 0-2/1-3) is then assigned for these contacts. (i.e. for contact-level 0 channels 0-1, 0-2 and 0-3 are averaged resulting in an average distance estimate equal to the distance between 0-2/1-3. For contact-level 1 channels 0-1, 1-2 and 1-3 are averaged and compared to the value of channel 0-2. This results in a power comparison between level 0 and 1 which is approximately considering the same (average) distance.) Furthermore, differences found in power are greater than those expected to be caused by the remaining small (average) difference in distance. This is confirmed by the results of an additional analysis showing that the predictive accuracy for the pattern based method when applying the DETEC or quadratic distance correction method resulted in similar results (84.6% accuracy without distance correction vs. 90.4% accuracy with distance correction using the "Max" feature on the "NL" data with clear beta activity).

For the "decision tree" technique, the differences in distance were accounted for in the structure of the tree during the design process. We additionally account for differences in distance by the fact that we iteratively narrow down the choices by looking at the second and third best options: contacts with bigger distances will require additional steps in the tree, that will narrow down the number of options. As a result, distance is intrinsically included in the choices for the branches. This is confirmed by the results of an additional analysis showing that the predictive accuracy for the decision tree method when applying the DETEC distance correction method resulted in similar results (86.5% accuracy without distance correction vs. 71.2% accuracy with distance correction using the "Max" feature on the "NL" data with clear beta activity.

**Supplementary figure 1.** Selection-trees for the online "decision tree" method.

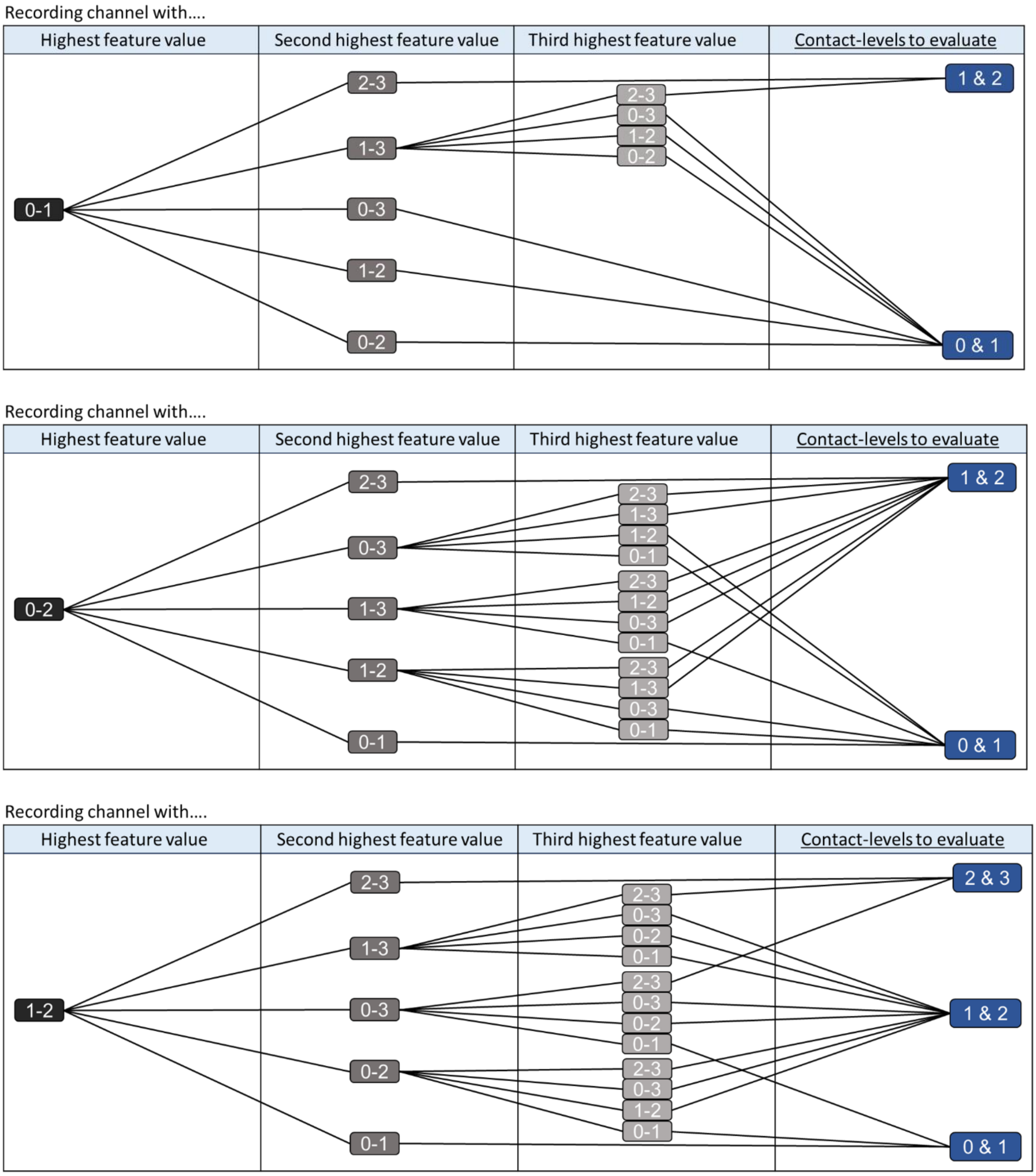

Recording channel with....

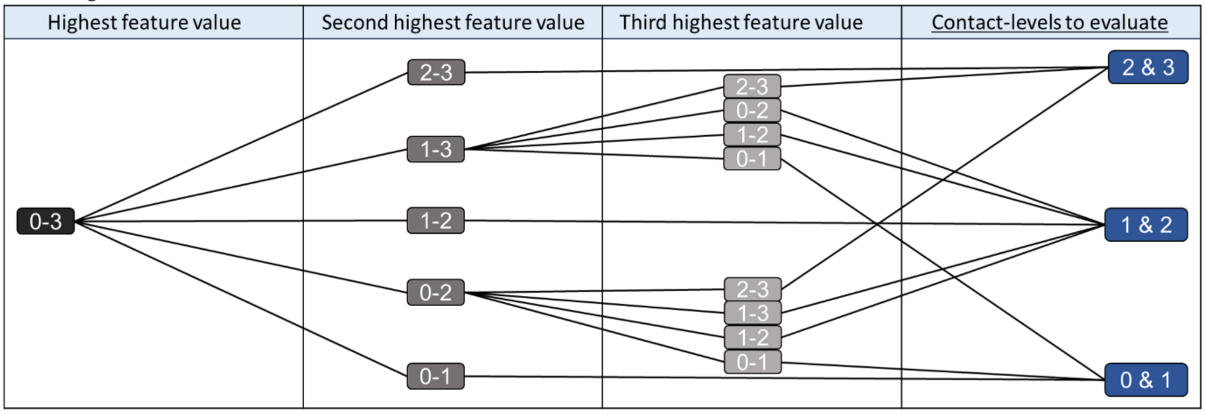


Recording channel with....

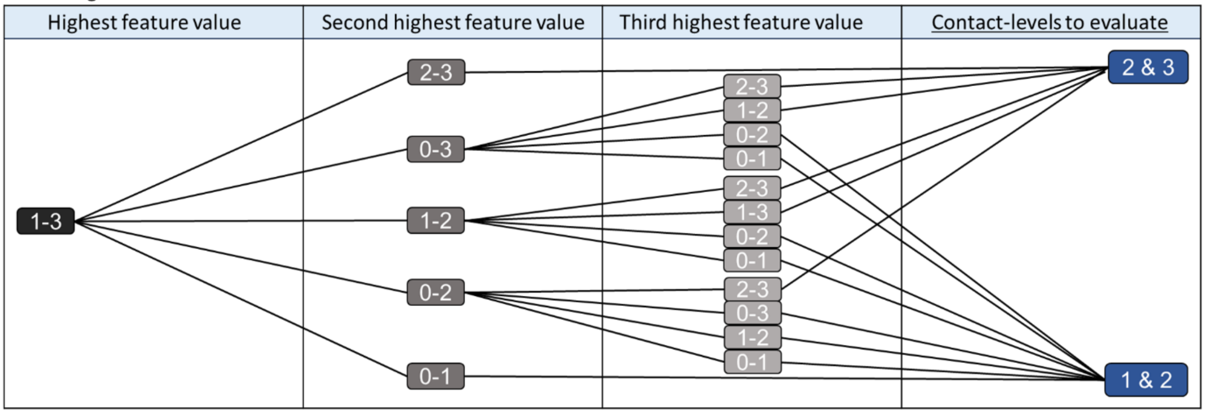


Recording channel with....

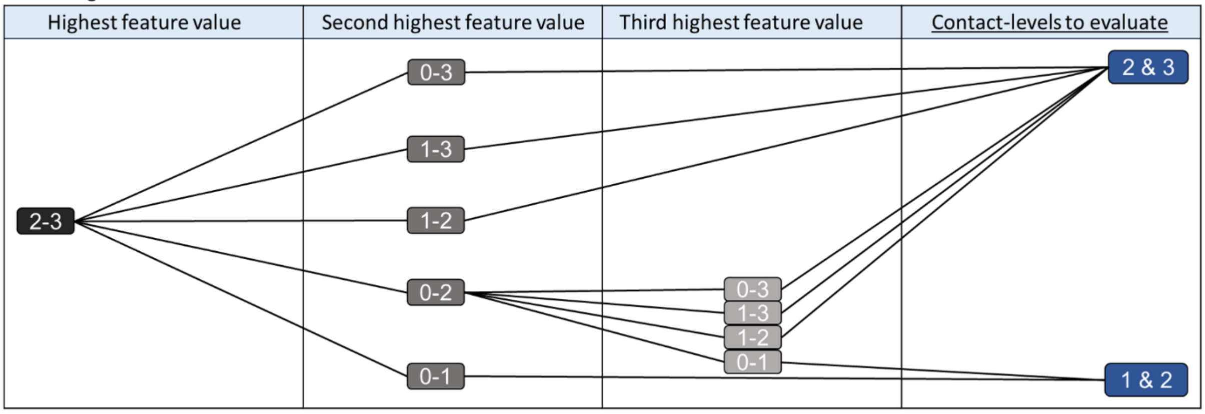

**Supplementary figure 2.** Elimination-trees for the online "decision tree" method.

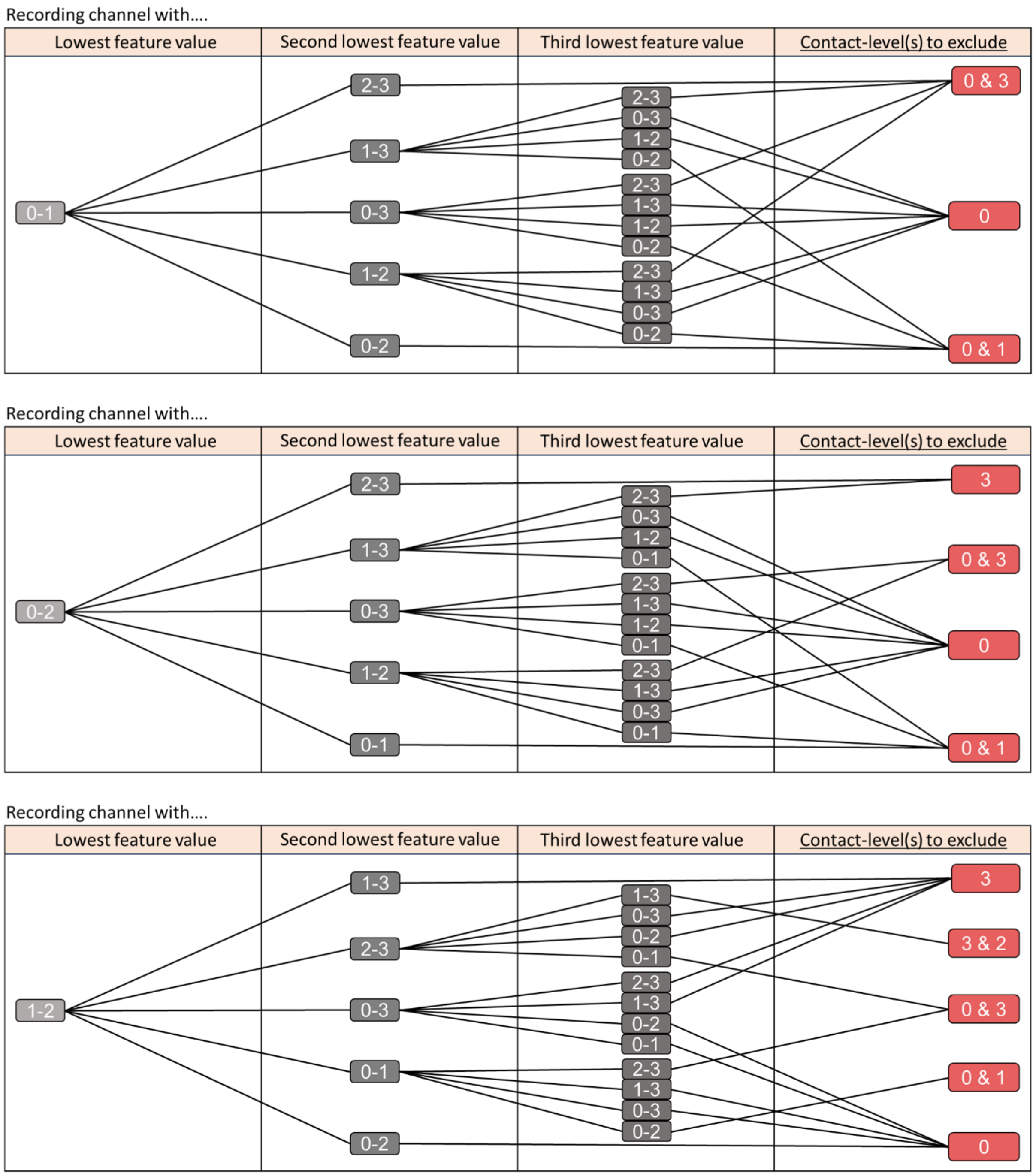

Recording channel with....

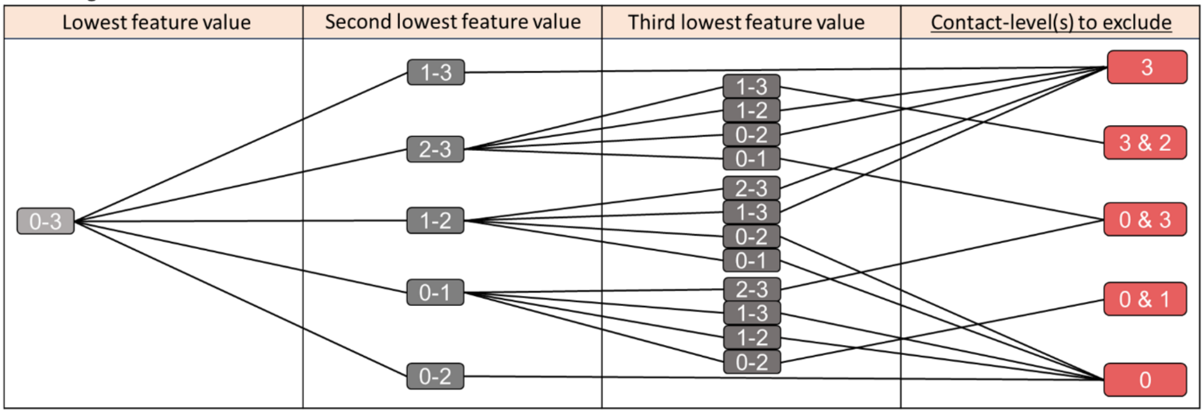


Recording channel with....

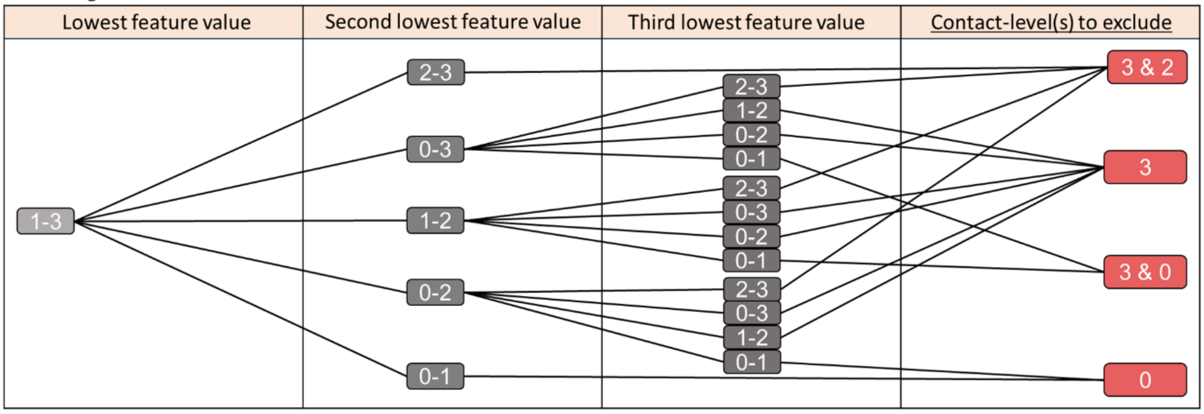


Recording channel with....

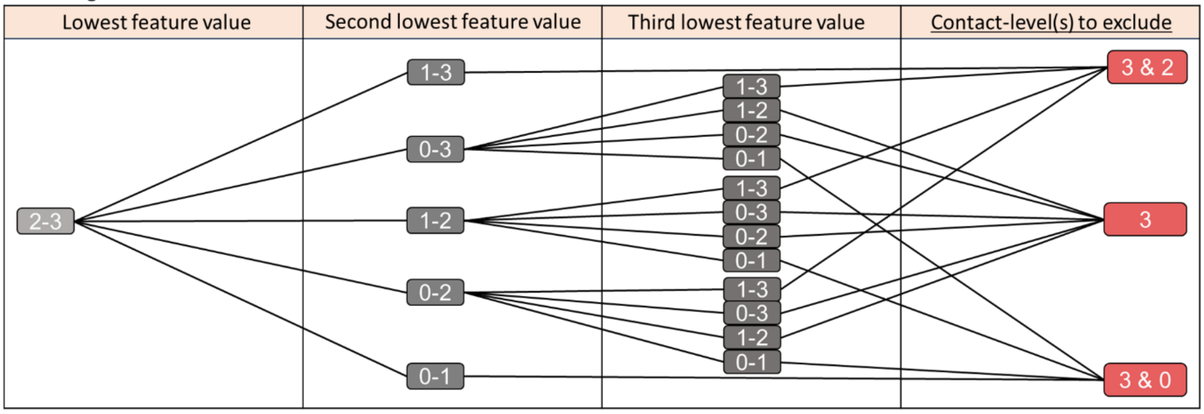

A

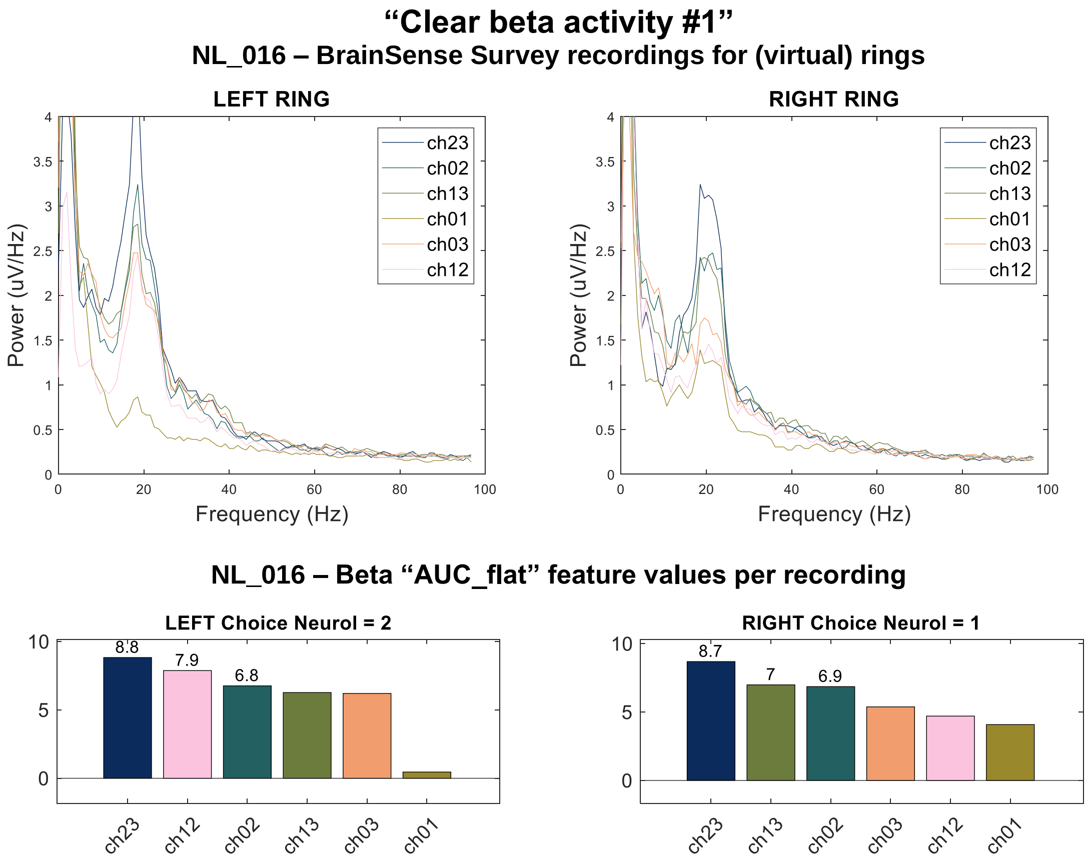

"Clear beta activity #1"
NL_016 – BrainSense Survey recordings for (virtual) rings
LEFT RING
RIGHT RING
Power (uV/Hz)
Frequency (Hz)
ch23
ch02
ch13
ch01
ch03
ch12
NL_016 – Beta "AUC_flat" feature values per recording
LEFT Choice Neurol = 2
RIGHT Choice Neurol = 1
8.8
7.9
6.8
8.7
7
6.9
ch23
ch12
ch02
ch13
ch03
ch01


B

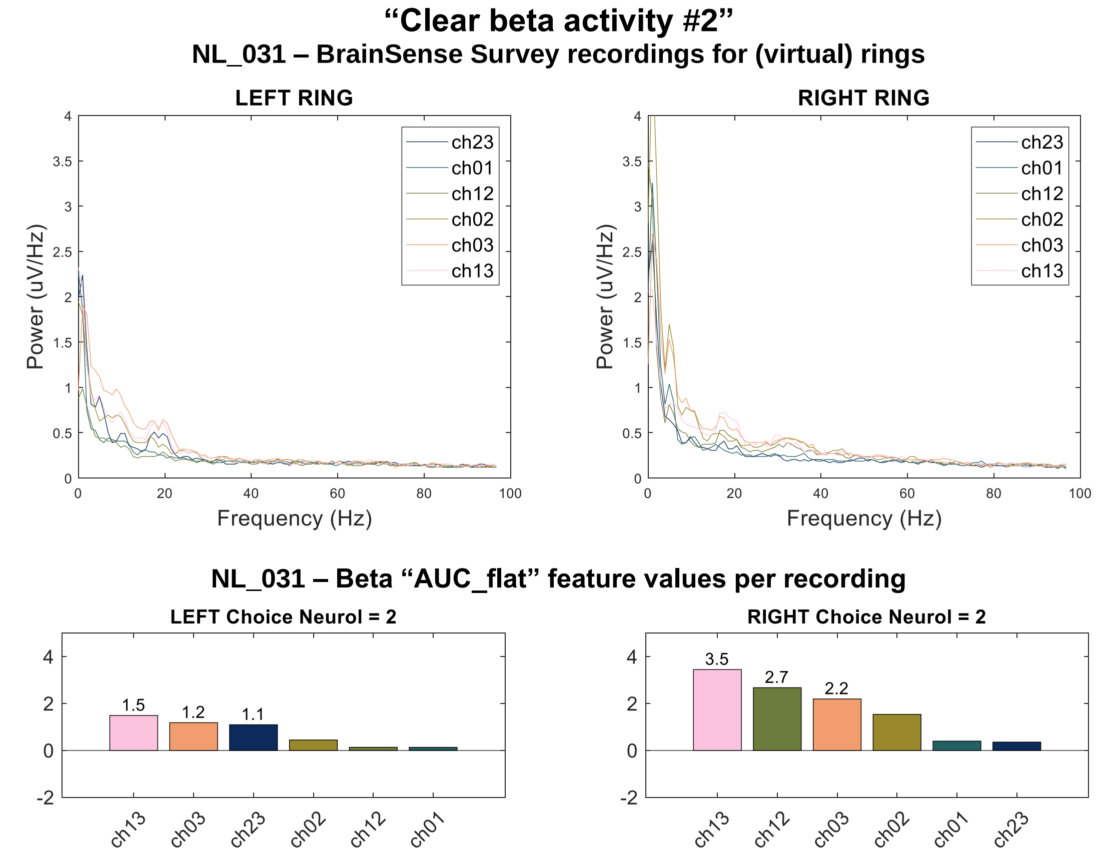

"Clear beta activity #2"
NL_031 – BrainSense Survey recordings for (virtual) rings
LEFT RING
RIGHT RING
Power (uV/Hz)
Frequency (Hz)
ch23
ch01
ch12
ch02
ch03
ch13
NL_031 – Beta "AUC_flat" feature values per recording
LEFT Choice Neurol = 2
RIGHT Choice Neurol = 2
1.5
1.2
1.1
3.5
2.7
2.2
ch13
ch03
ch23
ch02
ch12
ch01

C

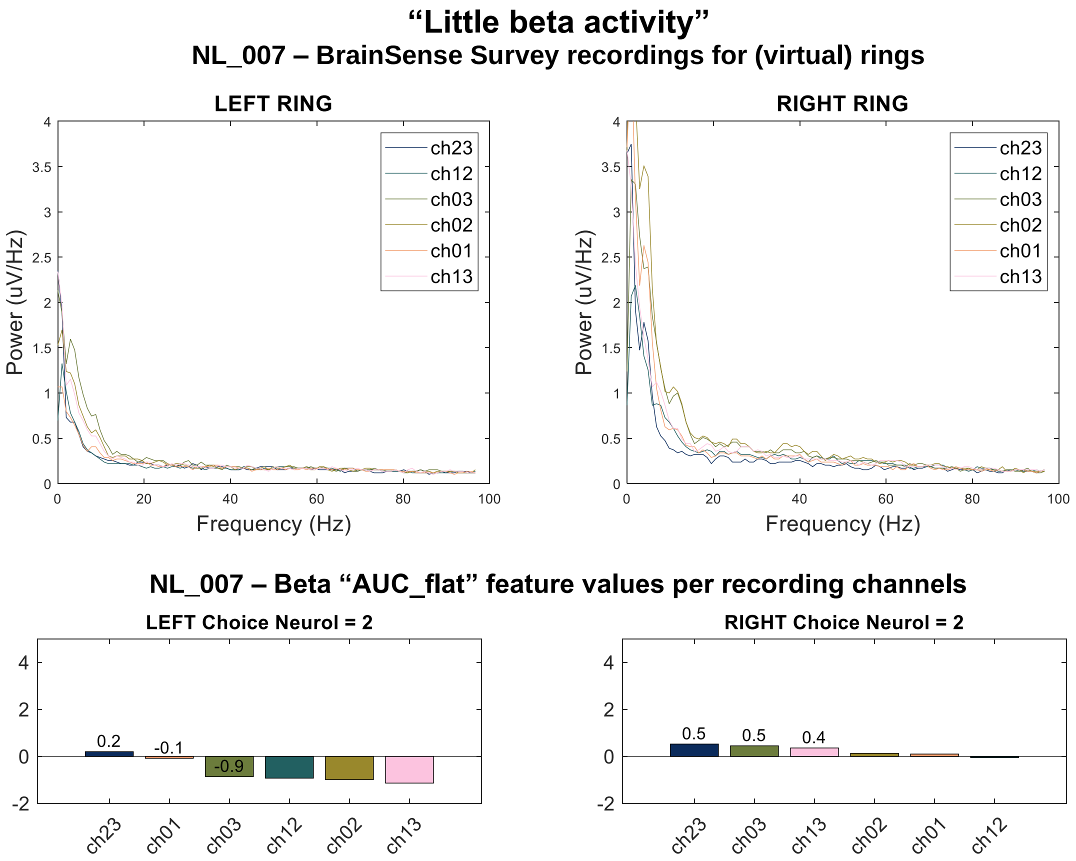

“Little beta activity”
NL_007 – BrainSense Survey recordings for (virtual) rings
LEFT RING
RIGHT RING
Power (uV/Hz)
Frequency (Hz)
ch23
ch12
ch03
ch02
ch01
ch13
NL_007 – Beta “AUC_flat” feature values per recording channels
LEFT Choice Neurol = 2
0.2
-0.1
-0.9
ch23
ch01
ch03
ch12
ch02
ch13
RIGHT Choice Neurol = 2
0.5
0.5
0.4
ch23
ch03
ch13
ch02
ch01
ch12


D

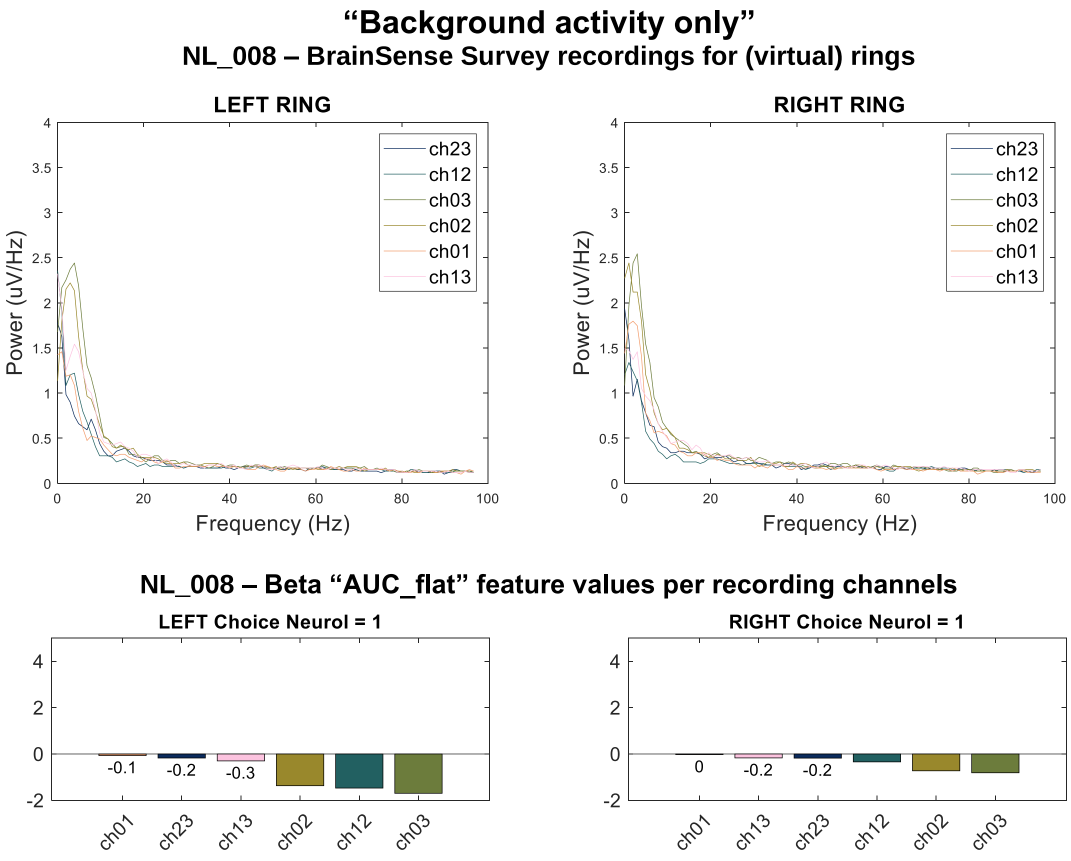

“Background activity only”
NL_008 – BrainSense Survey recordings for (virtual) rings
LEFT RING
RIGHT RING
Power (uV/Hz)
Frequency (Hz)
ch23
ch12
ch03
ch02
ch01
ch13
NL_008 – Beta “AUC_flat” feature values per recording channels
LEFT Choice Neurol = 1
-0.1
-0.2
-0.3
ch01
ch23
ch13
ch02
ch12
ch03
RIGHT Choice Neurol = 1
0
-0.2
-0.2
ch01
ch13
ch23
ch12
ch02
ch03

**Supplementary figure 3.** Individual cases within each of the three classes for the level of beta-activity: **A)** “clear beta activity – first example”: at least one channel with “AUC_flat” ≥ 0.6 μV, showing case with high clear beta activity; **B)** “clear beta activity – second example”: at least one channel with “AUC_flat” ≥ 0.6 μV, showing case with low clear beta activity; **C)** “little beta activity”: one or more channels with “AUC_flat” between 0.0 and 0.6 μV; and **D)** “background signal only”: all channels with “AUC_flat” ≤ 0.0 μV. Even in cases with a flattened power (“little beta” or “background signal only”), we could still extract an order from the global beta (i.e., background activity in the beta-band), as demonstrated by the “AUC_flat” feature value graphs.
