## Supplementary file 2 for "Online prediction of optimal deep brain stimulation contacts from local field potentials in chronically-implanted patients with Parkinson’s disease"

**Supplementary table 1.** Contact-level choices at monopolar review compared to chronic contact-level choices for all patients with measurements OFF-medication.

| Hemisphere | MPR contact | 6 month post-implant | 1 year post-implant |
|---|---|---|---|
| NL_007_L | 2 | 2 | 2 |
| NL_007_R | 2 | 2 | 2 |
| NL_008_L | 1 | 1 | 1 |
| NL_008_R | 1 | 1 | 1 |
| NL_016_L | 2 | 2 | 2 |
| NL_016_R | 1 | 1 | IL: 1 (70%) and 2 (30%) |
| NL_017_L | 1 | 1 | 3 |
| NL_017_R | 1 | 0 | 1 |
| NL_018_L | 2 | 2 | 2 |
| NL_018_R | 2 | 2 | 2 |
| NL_019_L | 1 | Not available | Not available |
| NL_019_R | 2 | Not available | Not available |
| NL_020_L | 2 | 2 | 2 |
| NL_020_R | 2 | 3 | 3 |
| NL_022_L | 2 | 2 | Not available |
| NL_022_R | 2 | 2 | Not available |
| NL_023_L | 2 | 2 | 2 |
| NL_023_R | 2 | 1 | 1 |
| NL_024_L | 2 | IL: 1 (50%) and 2 (50%) | IL: 1 (50%) and 2 (50%) |
| NL_024_R | 2 | 2 | 2 |
| NL_027_L | 1 | IL: 2 (40%) and 3 (60%) | IL: 2 (40%) and 3 (60%) |
| NL_027_R | 3 | 3 | 3 |
| NL_028_L | 2 | 2 | 2 |
| NL_028_R | 3 | 3 | 3 |
| NL_031_L | 2 | 2 | IL: 2 (20%) and 3 (80%) |
| NL_031_R | 2 | 1 | 2 |
| NL_037_L | 1 | 1 | 1 |
| NL_037_R | 1 | 1 | 1 |
| NL_042_L | 2 | 2 | 2 |
| NL_042_R | 2 | 2 | 2 |
| NL_044_L | 2 | 2 | 2 |
| NL_044_R | 2 | 2 | 2 |
| NL_045_L | 2 | Not available | Not available |
| NL_045_R | 2 | Not available | Not available |
| NL_046_L | 2 | Not available | Not available |
| NL_046_R | 2 | Not available | Not available |
| NL_048_L | 2 | 2 | 2 |
| NL_048_R | 2 | 2 | 2 |
| NL_049_L | 2 | 2 | 2 |
| NL_049_R | 1 | IL: 0 (50%) and 1 (50%) | IL: 0 (50%) and 1 (50%) |
| NL_051_L | 2 | 3 | 2 |
| NL_051_R | 1 | 2 | 2 |
| NL_052_L | 2 | 2 | 2 |
| NL_052_R | 2 | 2 | 2 |
| NL_053_L | 2 | 2 | 2 |

| | | | |
|---|---|---|---|
| NL_053_R | 2 | 2 | 2 |
| NL_054_L | 3 | Not available | Not available |
| NL_054_R | 2 | Not available | Not available |
| NL_055_L | 2 | 2 | 2 |
| NL_055_R | 2 | 2 | 2 |
| NL_056_L | 3 | 3 | 3 |
| NL_056_R | 2 | 2 | 2 |
| NL_057_L | 1 | 1 | 1 |
| NL_057_R | 1 | 1 | 1 |
| NL_058_L | 2 | 2 | 2 |
| NL_058_R | 2 | 2 | 2 |
| NL_059_L | 1 | 1 | 1 |
| NL_059_R | 2 | 2 | 2 |
| NL_065_L | 2 | Not available | Not available |
| NL_065_R | 2 | Not available | Not available |
| NL_068_L | 2 | 2 | Not reached |
| NL_068_R | 1 | 1 | Not reached |
| NL_069_L | 2 | 2 | Not reached |
| NL_069_R | 2 | 2 | Not reached |
| NL_070_L | 2 | 2 | Not reached |
| NL_070_R | 2 | 2 | Not reached |
| NL_071_L | 1 | 1 | Not reached |
| NL_071_R | 2 | 2 | Not reached |
| **Total NL** | **68** | **56** | **48** |
| **Total NL = 0** | **0 (0.0%)** | **1 (1.7%)** | **0 (0.0%)** |
| **Total NL = 1** | **16 (23.5%)** | **13 (22.4%)** | **11 (22.9%)** |
| **Total NL = 2** | **48 (70.6%)** | **36 (62.1%** | **30 (62.5%)** |
| **Total NL = 3** | **4 (5.9%)** | **6 (10.3%)** | **7 (14.6%)** |
| CH_101_L | 2 | - | 2 |
| CH_105_L | 1 | 1 | 2(b) |
| CH_106_L | 1 | - | 2 |
| CH_106_R | 1 | - | 2 |
| CH_110_R | 2 | - | 2(b) |
| CH_117_L | 2 | IL: 2 (50%) and 3 (50%) | IL: 2 (50%) and 3 (50%) |
| CH_117_R | 2 | 2 | 2 |
| CH_118_L | 1 | 1 | 1 |
| CH_118_R | 1 | 1 | 1 |
| CH_121_L | 2 | 2 | Not reached |
| CH_121_R | 2 | 2 | Not reached |
| CH_122_L | 1 | Not available | Not available |
| CH_122_R | 1 | Not available | Not available |
| CH_124_L | 1 | Not available | Not available |
| CH_124_R | 1 | Not available | Not available |
| CH_125_L | 1 | 1 | 1 |
| CH_125_R | 1 | 1 | 1 |
| CH_126_L | 2 | Not available | Not available |
| CH_126_R | 2 | Not available | Not available |
| CH_127_L | 1 | Not available | Not available |
| CH_127_R | 1 | Not available | Not available |
| **Total CH** | **21** | **9** | **11** |
| **Total CH = 0** | **0 (0.0%)** | **0 (0.0%)** | **0 (0.0%)** |
| **Total CH = 1** | **13 (61.9%)** | **5 (55.6%)** | **4 (36.4%)** |
| **Total CH = 2** | **8 (38.1%)** | **3.5 (38.9%)** | **6.5 (59.1%)** |
| **Total CH = 3** | **0 (0.0%)** | **0.5 (5.6%)** | **0.5 (4.5%)** |
| DE_009_L | 3 | IL: 1 (50%) and 2 (50%) | IL: 1 (50%) and 2 (50%) |

| | | | |
|---|---|---|---|
| DE_009_R | 0 | IL: 1 (50%) and 2 (50%) | IL: 1 (50%) and 2 (50%) |
| DE_010_L | 1 | 1 | 1 |
| DE_010_R | 2 | 2 | 2 |
| DE_011_L | 3 | 3 | 3 |
| DE_011_R | 2 | 2 | 2 |
| DE_012_L | 1 | explanted | explanted |
| DE_012_R | 0 | explanted | explanted |
| DE_013_L | 2 | 1 | 1 |
| DE_013_R | 0 | 1 | 1 |
| DE_015_L | 2 | 2 | 2 |
| DE_015_R | 2 | 3 | 2 |
| DE_016_L | 2 | IL: 1 (50%) and 2 (50%) | IL: 1 (50%) and 2 (50%) |
| DE_016_R | 2 | IL: 1 (50%) and 2 (50%) | IL: 1 (50%) and 2 (50%) |
| DE_017_L | 3 | 2 | 2 |
| DE_017_R | 3 | 2 | 2 |
| DE_018_L | 2 | IL: 1 (50%) and 2 (50%) | IL: 1 (50%) and 2 (50%) |
| DE_018_R | 2 | IL: 1 (50%) and 2 (50%) | IL: 1 (50%) and 2 (50%) |
| DE_019_L | 1 | 1 | IL: 1 (50%) and 2 (50%) |
| DE_019_R | 1 | IL: 1 (50%) and 2 (50%) | IL: 1 (50%) and 2 (50%) |
| DE_020_L | 1 | 1 | IL: 1 (50%) and 2 (50%) |
| DE_020_R | 0 | 0 | 0 |
| DE_024_L | 2 | 2 | 2 |
| DE_024_R | 2 | 2 | 2 |
| DE_026_L | 1 | IL: 1 (50%) and 2 (50%) | IL: 1 (50%) and 2 (50%) |
| DE_026_R | 1 | 2 | 2 |
| DE_033_L | 1 | 2 | explanted |
| DE_033_R | 1 | 1 | explanted |
| DE_035_L | 2 | 2 | not reached |
| DE_035_R | 1 | 1 | not reached |
| DE_036_L | 1 | 1 | not reached |
| DE_036_R | 1 | IL: 1 (50%) and 2 (50%) | not reached |
| **Total DE** | **32** | **30** | **24** |
| **Total DE = 0** | **4 (12.5%)** | **1 (3.3%)** | **1 (4.2%)** |
| **Total DE = 1** | **13 (40.6%)** | **12.5 (41.7%)** | **8 (33.3%)** |
| **Total DE = 2** | **11 (34.4%)** | **14.5 (48.3%)** | **14 (58.3%)** |
| **Total DE = 3** | **4 (12.5%)** | **2 (6.7%)** | **1 (4.2%)** |

Abbreviations MPR: monopolar review; IL: interleaved programming, Included contacts counted 0.5 times in IL programming; “Not available”: patients changed centres, therefore no recordings are available; “Not reached”: this follow-up time was not reached during the study period; “explanted”: leads were explanted during the study period.

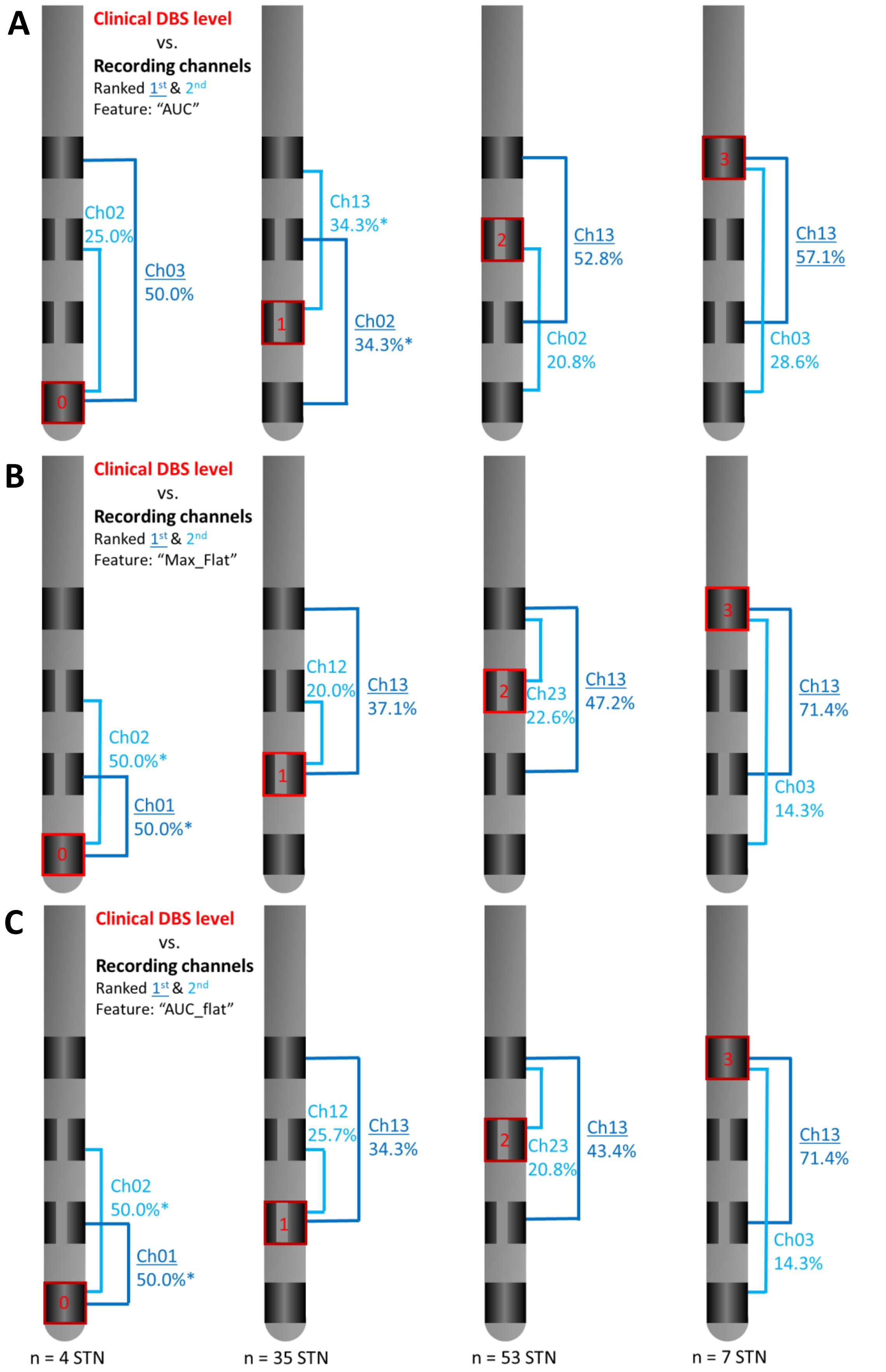


**Supplementary figure 1.** First- (dark blue) and second (light blue) ranked channel (Ch) per clinically chosen deep brain stimulation (DBS) contact-level at monopolar review (in red) for **a)** the area under the curve ("AUC"), **b)** the maximum power after removal of the aperiodic signal component ("Max_flat") and **c)** area under the curve after removal of the aperiodic signal component ("AUC_flat") beta-frequency band features. Showing how often the channel was ranked first based on this feature across the entire population of subthalamic nuclei (STN) as a percentage per 1st and 2nd ranked channel. Results shown are for all included leads with clear beta activity. (*) In case of an equal percentage (i.e. channels are ranked first with equal frequency) the channel which was ranked second most often, of these two options, was indicated as ranked 1st.

**Supplementary table 2.** Frequency of 1st/2nd ranked contact-level being the clinically chosen stimulation level for the selection only or selection and elimination decision tree models using all features (OFF-med.)

| | DT – "Max" | | DT – "AUC" | | DT – "Max_flat" | | DT – "AUC_flat" | |
|---|---|---|---|---|---|---|---|---|
| | **Sel. only** | **Sel. + El.** | **Sel. only** | **Sel. + El.** | **Sel. only** | **Sel. + El.** | **Sel. only** | **Sel. + El.** |
| **Training set "NL" (52 STN)** | 86.5% (45) | 88.5% (46) | 84.6% (44) | 88.5% (46) | 84.6% (44) | 86.5% (45) | 88.5% (46) | 88.5% (46) |
| **Validation set "CH" (15 STN)** | 86.7% (13) | 93.3% (14) | 86.7% (13) | 93.3% (14) | 53.3% (8) | 60.0% (9) | 60.0% (9) | 73.3% (11) |
| **Validation set "DE" (32 STN)** | 75.0% (24) | 78.1% (25) | 78.1% (25) | 84.4% (27) | 84.4% (27) | 87.5% (28) | 78.1% (25) | 78.1% (25) |

Abbreviations: DT: Decision tree; Max: maximum power feature; AUC: area under the receiver operator curve feature; _flat: after removing aperiodic signal component (1/frequency); Sel. only: selection tree only; Sel. + El.: selection and elimination trees combined

**Supplementary table 3.** Frequency of 1st–only and 1st/2nd ranked contact-level corresponding to the clinically chosen stimulation level for the pattern based model ("Max" feature) and DETEC algorithm.†

| | 1st ranked only | | 1st / 2nd ranked | |
|---|---|---|---|---|
| | **Pattern based – "Max"** | **DETEC algorithm** | **Pattern based – "Max"** | **DETEC algorithm** |
| **Training set "NL"** (52 STN) | 73.1% (38) | 15.4% (8) | 84.6% (44) | 63.5% (33) |
| **Validation set "CH"** (15 STN) | 40.0% (6) | 26.7% (4) | 66.7% (10) | 46.7% (7) |
| **Validation set "DE"** (32 STN) | 56.3% (18) | 25.0% (8) | 71.9% (23) | 65.6% (21) |

† Based on OFF medication state LFP recording data only
Abbreviations: Max: maximum power feature

**Supplementary table 4.** Frequency of 1st/2nd ranked contact-level corresponding to the clinically chosen stimulation level for the decision tree and pattern based models ("Max" feature) for "NL" patients when considering low- and high-beta individually.†

| | Low-beta (13-20Hz) | | High-beta (21-35Hz) | |
|---|---|---|---|---|
| | **Decision tree* – "Max"** | **Pattern based – "Max"** | **Decision tree* – "Max"** | **Pattern based – "Max"** |
| **Training set "NL"** (52 STN) | 86.5% (45) | 84.6% (44) | 88.5% (46) | 84.6% (44) |

* Results for selection decision tree only.
† Based on OFF medication state LFP recording data only
Abbreviations: Max: maximum power feature

| **Supplementary table 5.** Frequency of 1st/2nd ranked contact-level being the clinically chosen stimulation level per ranking model and all available features for all three centres (OFF-medication state only). | | | | | | | | |
|---|---|---|---|---|---|---|---|---|
| | **Decision tree* (1st / 2nd ranked)** | | | | **Pattern based (1st / 2nd ranked)** | | | |
| | **Max** | **AUC** | **Max_flat** | **AUC_flat** | **Max** | **AUC** | **Max_flat** | **AUC_flat** |
| **Training set "NL" (52 STN)** | 86.5% (45) | 84.6% (44) | 84.6% (44) | 88.5% (46) | 84.6% (44) | 82.7% (43) | 90.4% (47) | 90.4% (47) |
| **Validation set "CH" (15 STN)** | 86.7% (13) | 86.7% (13) | 53.3% (8) | 60.0% (9) | 66.7% (10) | 80.0% (12) | 66.7% (10) | 66.7% (10) |
| **Validation set "DE" (32 STN)** | 75.0% (24) | 78.1% (25) | 84.4% (27) | 78.1% (25) | 71.9% (23) | 68.8% (22) | 78.1% (25) | 81.3% (26) |

* Results for selection decision tree only.
Abbreviations: Max: maximum power feature; AUC: area under the receiver operator curve feature; _flat: after removing aperiodic signal component (1/frequency)

| **Supplementary table 6.** Patient characteristics for subset of "NL" patients with and without stun effect | | | |
|---|---|---|---|
| | **"NL" stun effect subset (n=17)** | **"NL" no stun effect subset (n=24 (7 duplicate))** | **P-value†** |
| Age at surgery (years) | 63.2 (SD 8.3) | 62.8 (SD 7.7) | 0.701 |
| Male, N (%) | 12 (70.6%) | 15 (62.5%) | 0.591 |
| Disease duration at recording (years) | 9.8 (SD 3.6) | 9.7 (SD 4.4) | 0.841 |
| Clinical total pre-operative OFF-med motor score | 34.8 (SD 11.7) | 42.8 (SD 11.1) | **0.029** |
| Clinical total pre-operative ON-med motor score | 15.3 (SD 8.4) | 20.7 (SD 8.9) | 0.059 |
| Levodopa response *(OFF-ON/OFF*100)* | 55.9 (SD 19.8) | 51.8 (SD 17.9) | 0.317 |
| Preoperative LEDD | 1176 (SD 550) | 1403 (SD 567) | 0.209 |
| Time since lead implantation at LFP (days) | 8.7 (SD 2.4) | 7.8 (SD 3.2) | 0.365 |
| Time since lead implantation at MPR (days) | 9.3 (SD 1.0) | 9.2 (SD 1.1) | 0.742 |

† Mann-Whitney- U test (numerical data); Chi-square test (categorical data).
* Significant difference between groups (two-sided p-value < 0.05).
Abbreviations: ON-med: ON-medication; n: number of patients; SD: standard deviation; DBS: deep brain stimulation; LEDD: levodopa equivalent daily dose.

| **Supplementary table 7.** Patient characteristics for subset of “CH” patients OFF- vs. ON-medication | | | |
|---|---|---|---|
| | **“CH” OFF-med subset (n=12)** | **“CH” ON-med subset (n=18)** | **P-value†** |
| Age at surgery (years) | 64.4 (SD 6.0) | 64.6 (SD 7.8) | 0.687 |
| Male, N (%) | 9 (75.0%) | 9 (50.0%) | 0.171 |
| Disease duration at recording (years) | 9.1 (SD 2.8) | 10.3 (SD 4.6) | 0.653 |
| Clinical total pre-operative OFF-med motor score | 39.7 (SD 11.4) | 41.7 (SD 22.4) | 0.689 |
| Clinical total pre-operative ON-med motor score | 14.4 (SD 8.2) | 18.9 (SD 23.4) | 0.843 |
| Levodopa response *(OFF-ON/OFF*100)* | 66.1% (SD 15.8%) | 62.8% (19.9%) | 0.644 |
| Preoperative LEDD | 1112 (SD 583) | 1104 (SD 568) | 1.000 |
| Time since lead implantation at LFP (days) | 5.6 (SD 1.5) | 12.4 (SD 30.2) | 0.621 |
| Time since lead implantation at MPR (days) | 20.8 (SD 32.3) | 30.1 (SD 38.3) | 0.488 |

† Mann-Whitney U test (numerical data); Chi-square test (categorical data).
* Significant difference between groups (two-sided p-value < 0.05).
Abbreviations: ON-med: ON-medication; n: number of patients; SD: standard deviation; DBS: deep brain stimulation; LEDD: levodopa equivalent daily dose.

**Supplementary table 8.** Contact-level choices at monopolar review compared to chronic contact-level choices for all "CH" patients with measurements ON-medication.

| Hemisphere | MPR contact | 6 month post-implant | 1 year post-implant |
|---|---|---|---|
| CH_102_L | 2 | - | 2 |
| CH_103_L | 1 | Not available | Not available |
| CH_103_R | 1 | Not available | Not available |
| CH_104_L | 2 | - | 2 |
| CH_104_R | 2 | - | 2 |
| CH_109_L | 1 | Not available | Not available |
| CH_109_R | 1 | Not available | Not available |
| CH_111_L | 2 | - | IL: 2 (50%) and 3 (50%) |
| CH_111_R | 2 | - | 1(b) |
| CH_112_L | 2 | IL: 1 (50%) and 2 (50%) | IL: 1 (50%) and 2 (50%) |
| CH_112_R | 2 | 2 | 2 |
| CH_113_L | 2 | IL: 2 (50%) and 3 (50%) | - |
| CH_113_R | 2 | 2 | - |
| CH_114_L | 2 | 2(a) | 2(a) |
| CH_114_R | 2 | 2 | 2 |
| CH_115_L | 2 | Not available | Not available |
| CH_115_R | 2 | Not available | Not available |
| CH_116_L | 2 | Not available | Not available |
| CH_116_R | 2 | Not available | Not available |
| CH_119_L | 1 | 1 | - |
| CH_119_R | 1 | 1 | - |
| CH_120_L | 3 | 3 | 3 |
| CH_120_R | 1 | 1 | 1 |
| CH_123_L | 1 | 1 | - |
| CH_123_R | 1 | 1 | - |
| CH_128_L | 2 | Not available | Not available |
| CH_128_R | 1 | Not available | Not available |
| CH_129_L | 2 | Not available | Not available |
| CH_129_R | 2 | Not available | Not available |
| CH_130_L | 2 | 2 | - |
| CH_130_R | 2 | IL: 1 (50%) and 2 (50%) | - |
| CH_131_L | 2 | Not available | Not available |
| CH_131_R | 2 | Not available | Not available |
| CH_132_L | 2 | Not available | Not available |
| CH_132_R | 1 | Not available | Not available |
| **Total CH** | **35** | **13** | **11** |
| **Total CH = 0** | **0 (0.0%)** | **0 (0.0%)** | **0 (0.0%)** |
| **Total CH = 1** | **11 (31.4%)** | **6 (46.2%)** | **2.5 (22.7%)** |
| **Total CH = 2** | **23 (65.7%)** | **6.5 (50.0%)** | **7 (63.6%)** |
| **Total CH = 3** | **1 (2.9%)** | **0.5 (3.8%)** | **1.5 (13.6%)** |

Abbreviations MPR: monopolar review; IL: interleaved programming; "-": No recording at this time point; "Not available": patients changed centres, therefore no recordings are available
